# Quantifying Sleep-Wake Rhythms in the Hospital Environment with Digital Technologies

**DOI:** 10.1101/2025.11.18.25340421

**Authors:** Carsten Skarke, Nadim El Jamal, Michael V. Genuardi, Thomas G. Brooks, Antonijo Mrčela, Nicholas F. Lahens, Makayla Cordoza, Sean Sarles, Amarachi Mbadugha, Dineth R. Karunamuni, Shikhar Gupta, Charles J. Bae, Amita Sehgal, Thomas P. Cappola, Jacob T. Gutsche, Pavan Atluri, Nimesh D. Desai, Michael A. Acker, Gregory R. Grant, Richard J. Schwab, Ilene M. Rosen, Garret A. FitzGerald

**Affiliations:** Institute for Translational Medicine and Therapeutics (ITMAT), University of Pennsylvania Perelman School of Medicine, Philadelphia, PA 19104, USA; Department of Medicine, University of Pennsylvania Perelman School of Medicine, Philadelphia, PA 19104, USA; Department of Anesthesiology and Critical Care, University of Pennsylvania Perelman School of Medicine, Philadelphia, PA 19104, USA; Department of Surgery, University of Pennsylvania Perelman School of Medicine, Philadelphia, PA 19104, USA; School of Nursing, Vanderbilt University, Nashville, TN 37240, USA; Department of Genetics, University of Pennsylvania Perelman School of Medicine, Philadelphia, PA 19104, USA

## Abstract

Postoperative clinical care is prone to circadian desynchronization that, in turn, may influence health outcomes. We collected 1.8 million data points using 11 remote sensors during preoperative, in-hospital and post-discharge settings in 13 elective cardiac surgery patients. We found that room traffic continued during nighttime with ≥1 visit/h. Sound levels exceeded the recommended 45 dBA threshold (51.9±3.3 versus 48.3±4.2 dBA during nighttime). Brightness dropped at night (89.9±87.7 to 3.7±9.8 lux), but bright light exposures occurred. Ambient room temperature lacked sleep-inducing diurnal variability. Behavioral-physiological rhythms were disrupted (decreased amplitude of heart rate variability; *unadjusted-p*-*value*=0.03) and phase-shifted during hospitalization. Time awake during night hours increased from 10.7±7.9% preoperatively to 34.8±29.1% in-hospital (*unadjusted p-value=*0.0098). Cognitive function scores decreased (26.8±2.8 points preoperatively to 24.7±3.9 points in-hospital) with 31% of patients developing transient mild impairment. These data will inform the design of a controlled trial seeking to modify circadian/diurnal disruptors to enhance patient outcomes.

## Introduction

The hospital environment is an intensively monitored clinical setting where vital signs including heart rate, blood pressure, respiratory rate, temperature, and oxygen saturation are frequently tracked alongside other physiological parameters such as fluid balance and nutritional status. This comprehensive monitoring enables early detection of clinical deterioration and supports timely therapeutic intervention. Yet, for patients, the same 24-hour care introduces major challenges for maintaining normal sleep-wake cycles and alignment of internal biological rhythms with the external day-night cycle.

Sleep disruption constitutes one of the most common and distressing patient experiences during hospitalization ^1^, which, confirmed by polysomnographic recordings showing profound fragmentation of sleep architecture ^2^, is often precipitated by medications, comorbidities, nursing interventions and surgical procedures ^3^. In acute and critically ill patients, desynchronized sleep-wake rhythms have been linked to neurocognitive disturbances ^4^, affecting up to 25% of postoperative patients and as many as 80% of older adults ^5^. These adverse effects carry broad public health implications. Estimates of the financial burden range from tens of thousands annually per patient in the United States to hundreds of billions annually across U.S. healthcare systems due to longer hospital stays, outpatient visits, nursing home care, and rehabilitation ^6,7^.

Despite the rich data streams already generated in hospital settings, comprehensive assessments of patients’ biological rhythms and their relationship to postoperative outcomes remain understudied ^8^. Efforts to date largely center on critical care settings where desynchronization found, for example, in core body temperature ^9^, blood pressure ^10^ and heart rate ^11^ has been associated with worse clinical outcomes.

Digital medical approaches have the potential to expand patient risk assessment beyond the in-hospital monitoring to include preoperative and post-discharge health monitoring. Many of these platforms enable high-resolution sampling to examine diurnal variance more closely and offer healthcare strategies to deliver scalable, adaptive, and patient-centered solutions.

In this investigation, we present a pilot multimodal characterization of behavioral, physiological and sleep-wake rhythms of patients undergoing elective cardiac surgery using comprehensive digital monitoring, and to pair this with assessments of cognitive function. We will use this information to address the hypothesis that a reduction in circadian disruption leads to improved clinical outcomes.

## Methods

### Patients

Our study consented (per IRB protocol #852451 approved on November 21, 2022, ClinicalTrials.gov Identifier: NCT05828680 registered on April 24, 2023) and enrolled adult patients scheduled at the Hospital of the University of Pennsylvania for elective cardiac surgery procedures of coronary artery bypass, valve repairs or replacements, and aortic root and hemiarch repair or replacement (May 2023 to July 2024). We excluded patients with reported or suspected sleep disturbances such as sleep apnea, who took sleep aids or sedating medications, with substance use disorders, with hearing deficits to minimize discomfort from the in-ear vital sign device, (c-med alpha, Cosinuss GmbH, München, Germany), any skin concerns on device placement locations, or who had a recent seizure. For mobile phone app compatibility, we excluded patients who did not use iPhones.

### Study procedures

Between 7 and 14 days before the scheduled surgery, patients were asked to continuously wear a wrist actigraphy device (AX3 with wristband, Axivity, Newcastle upon Tyne, UK) for 7 days and apply an electrocardiogram (ECG) (Bittium Faros 180, Bittium Corporation, Oulu, Finland) for 48 hours. They were asked to perform the phone-based psychomotor vigilance test (PVT; NASA PVT+, Fatigue Countermeasures Laboratory, NASA Ames Research Center, Moffett Field, CA, USA) in the morning, noon, and evening for 7 days and were prompted to answer surveys using the Beiwe app (Onnela Lab, Harvard Chan School of Public Health, Boston, MA, USA) assessing their mood. The Beiwe app also collected passive data in an anonymized manner such as mobility patterns, mood, and pain level ^12^. A certified study coordinator administered the paper version 8.1 of the Montreal Cognitive Assessment (MoCA, certificate of completion USBANLA710659639-01 acknowledged to LaVenia Banas on June 14, 2023). Study procedures were then halted until their surgery. The typical postoperative course for cardiac surgery patients at the Hospital of the University of Pennsylvania was transfer from the operating room to the cardiac intensive care unit followed by a transfer to the cardiac step-down unit (SDU) and then discharged home. Study procedures were resumed the day after a patient was transferred to the SDU and consisted of patient centered and environment centered monitoring. In the SDU, ECG and limited-channel polysomnography (PSG; Prodigy Sleep System, Cerebra Medical Ltd., Winnipeg, Manitoba, CAN) were applied for 48 hours. Cerebra PSG devices were exchanged every 12 hours due to battery life constraints. The patients were also asked to wear the wrist actigraphy device, perform the mobile phone-based psychomotor vigilance test and answer the Beiwe surveys for the remainder of the stay. The hospital environment centered devices monitored light intensity and spectrum (XL-500 light spectrophotometer, nanoLambda, Daejeon, ROK), temperature and relative humidity (RH; HOBO Temperature/Relative Humidity/Light data logger, Onset, Bourne, MA, USA), and sound intensity (Class/Type 1 sound level meter, Quest™ Sound Examiner SE-401, TSI Incorporated, Shoreview, MN, USA), all custom mounted on three hospital-grade channel mounts (GCX Corporation, Petaluma, CA, USA). We coined this as the “Chronobiome Weather Station” in reference to our ongoing work on the human chronobiome ^13^ (***Supplementary Figure 1***). In addition, Pearl People Counter devices (SMS Store Traffic, St-Jean-Sur-Richelieu, Quebec, CAN), that record the amount of traffic into and out of the patient’s room, were mounted onto the metal frame of the entrance door (***Supplementary Figure 1***). An observer was also stationed outside the patient’s door to record the source of sounds the patient would hear and the number and purpose of traffic into the patient’s room. The environment centered devices were placed for a period of 48 hours, beginning at the postoperative resumption of study procedures. At that time, a second paper version (8.2 to prevent learning effects) of the MoCA test and the 3D-CAM ^14^ (3-minute diagnostic interview for CAM-defined delirium) were administered. The patients continued wearing the actigraphy device, answering Beiwe surveys, and performing the PVT until 7 days post-discharge. At 7 days post-hospital discharge, patients repeated the first paper version of the MoCA (8.1).

### Analysis Metrics

#### Data collected from patient room sensors (nearables)

Data from the Pearl People Counter were summed up to hourly totals across the 24-hour day. To validate against observer recordings, observer data were summed in the same method, and all measurements were compared by fitting a linear regression model (observer data vs Pearl People Counter data) and extracting the R-squared statistic. The Class/Type 1 sound level meter reported a-weighted Leq-1(Equivalent sound level over 1 second) every 5 seconds. These were binned into hourly means for our analyses. The nano lambda light spectrophotometer provided recordings every 11 seconds. These measurements were binned by taking the maximum recordings per wavelength per hour.

#### Data collected from patient-worn sensors (wearables)

The Cerebra PSG provided European data format files that were scored by a trained PSG technologist. The device also provided proprietary Odds Ratio Product values (ORP) that assess sleep depth ^15,16^ by reporting the relationships of the powers of different EEG frequencies in a single index ranging from zero reflecting very deep sleep to 2.5 indicating full wakefulness.

We set the activity accelerometer to measure acceleration in three axes at a rate of 100Hz. The data were then submitted to a machine learning pipeline (https://github.com/tgbrooks/biobankAccelerometerAnalysis) that resulted in an acceleration vector magnitude adjusted for gravity and a sleep probability (value between 0 and 1) every 30 seconds ^17^. To achieve a higher degree of confidence in sleep calls, we considered a probability of 0 as a wake call and a probability of 1 as a sleep call and eliminated the minority of values in between (10%, i.e. 54,792 out of 519,751 values). We validated the machine learning sleep and wake assessments against the PSG technologist’s readings by calculating the area under (AUC) the receiver operator characteristic (ROC) curve using the ROC and AUC functions from the R (v.4.3.2-4.5.1) package pROC (v.1.19.0.1)

To assess diurnal variability in activity levels we used the vector magnitude acceleration measurements to fit COSINOR curves and extract rhythmic parameters of amplitude, phase, and MESOR ^18^.

Heart rate variability (HRV) metrics were computed from the Bittium ECG data using Kubios HRV Scientific (version 4.1.0, Kubios, Kuopio, Finland). This software includes preprocessing of HRV data consisting of automatic noise detection and correction of ectopic, misplaced and artifact beats to enable analysis of long-term ambulatory recordings where signal quality varies (as we implemented previously ^19^). The resulting metrics were also used to assess diurnal variability by plotting COSINOR curves.

### Statistical Analyses

For statistical comparisons, we used paired Wilcoxon rank sum tests (as implemented by the wilcox.test function in R through the ggpubr package (v0.6.1) when n≥5 repeat measurements were present per group. For acrophase comparisons we used Kuiper tests from the ‘twosamples’ R package (v2.0.1) to account for the circular nature of acrophase. Because of the strong association between age and cognitive performance, we used age adjusted mixed effects linear regression models (lmer function from the lmerTest package in R, v3.1-3) to assess the effects the hospitalization and surgery had on MoCA scores. We report unadjusted *p*-values that are not corrected for multiple testing.

## Results

### Patient Cohort

Review of the operating schedule identified n=164 eligible patients per protocol inclusion and exclusion criteria specified in the methods (***Supplementary Figure 2***). A cohort of n=14 consented to participate. Reasons to decline included “too busy” and “already enough stress”. Data were collected on n=13, 54% female, with a mean±SD age of 61.6±8.7 years at enrollment scheduled for elective cardiovascular surgery for aortic valve and ascending aorta replacement (n=3), minimally invasive mitral valve replacement (n=3), aortic valve replacement (n=3) and other procedures as listed in ***Table 1***. Most surgeries were conducted in the morning hours, with only two patients operated on in the afternoon. Average hospital length of stay was 7.4±3.0 days, of which 5.8±3.1 days were in the cardiac surgery Step Down Unit (SDU). During SDU admission, most patients were on combination therapies of sedatives/analgesics (12/13), antihypertensives/diuretics (11/13), and antiarrhythmics/betablockers (10/13). Only two patients were on melatonin and diphenhydramine (***Supplementary Table 1***).

**Table 1.** Demographics.

| Age Range*<br>[yrs at enrollment] | Sex | Height<br>[cm] | Weight<br>[kg] | Elective surgery | Time of surgery | Time in<br>Hospital [d] | Time in SDU<br>[d] |
| --- | --- | --- | --- | --- | --- | --- | --- |
| 56-60 | M | 185 | 94.3 | AV and Ascending Aorta Repl. | Morning | 5.2 | 4.3 |
| 61-65 | F | 160 | 93.9 | AV and Ascending Aorta Repl. | Morning | 5.2 | 4.2 |
| 51-55 | F | 180 | 84.4 | Minimally invasive MVR | Morning | 15.4 | 14.2 |
| 56-60 | F | 167 | 75.8 | Minimally invasive MVR | Morning | 6.4 | 4.9 |
| 61-65 | F | 163 | 146.3 | AVR | Afternoon | 4.0 | 1.9 |
| 66-70 | F | 155 | 61.4 | Minimally invasive MVR | Morning | 8.2 | 6.9 |
| 61-65 | M | 157 | 73 | Ascending aorta graft | Morning | 6.2 | 4.9 |
| 66-70 | M | 183 | 83 | AV and Ascending Aorta Repl. | Morning | 8.1 | 6.9 |
| 56-60 | M | 178 | 100 | Redo-MVR | Afternoon | 9.9 | 8.7 |
| 61-65 | F | 135 | 73 | AVR | Morning | 7.3 | 6.1 |
| 36-40 | F | 150 | 69 | ROSS | Morning | 9.3 | 2.7 |
| 71-75 | M | 180 | 104.3 | AVR | Morning | 6.1 | 5.3 |
| 71-75 | M | 173 | 116 | CAB | Morning | 5.2 | 4.3 |

\* 36-40, 41-45, 46-50, 51-55, 56-60, 61-65, 66-70, 71-75 yrs

### Cardiac Surgery Patient Phenotypes Quantified in the Hospital Environment

We collected 1.8 million data points across 160 features during the preoperative, in-hospital and post-discharge collection periods (***Supplementary Figure 3***, ***Supplementary Table 2***). Self-reported levels of pain assessed daily through a phone app increased postoperatively by 210% compared to at home assessments during the preoperative phase. On average (±SD), patients started to report pain scores 2.8±1.3 days after operation coinciding with their transfer from ICU to SDU. The normalized (by the pre-operation, in-hospital or post-operation observation duration) the pain-score AUCs changed from 1.34±0.97 NRS (0-10 Numeric Rating Scale) preoperative, to 4.15±1.63 NRS in-hospital (*unadjusted p-value=*0.031 paired Wilcoxon rank sum test) and rebounded to 2.78±1.41 NRS after discharge. Similarly, self-reported fatigue in-hospital increased by 113% from the preoperative baseline. Here, the normalized AUCs were overall higher with 2.91±1.79 NRS pre-op, 6.21±0.9 NRS in-hospital (*unadjusted p-value=*0.031 paired Wilcoxon rank sum test) and 4.58±1.27 NRS post-discharge (***Supplementary Figure 4***). The emotional valence collected through phone-based surveys reflected the change in feelings between home and hospitalization with increased normalized AUCs of feeling blue (by 54%) and a corresponding loss in happiness (by 33%) (*unadjusted p-value=*0.031 paired Wilcoxon rank sum test) (***Supplementary Figure 5***, ***Supplementary Table 3***).

Phone-based geolocation data showed the extent to which admission to the hospital room disrupted daily routines. Here, physical diurnal rhythms dropped by 58.1% from a normalized AUC of 0.43±0.28 consistency degree (Interval [Int 0,1]) preoperatively to 0.18±0.08 Int in-hospital and consolidated to 0.3±0.13 Int at home (***Supplementary Figure 6***). The normalized AUC of phone app-based accelerometry decreased by 88% from 128.1±151.6 milli-g-force (normalized by the total observation duration) pre-operatively to 14.7±23.3 milli-g-force in-hospital (*unadjusted p-value=*0.047 paired Wilcoxon rank sum test).

Turning to physical activity measured by a wrist actigraphy device, we found that the rhythm-adjusted mean, the MESOR, declined post-operatively by 41.6% (median [interquartile range or IQR] 7.2[2.3] milli-g-force) compared to the pre-operative condition (17.3[9.0] milli-g-force, *unadjusted p-value=*0.002 paired Wilcoxon rank sum test) and recovered after discharge (11.0[4.7] milli-g-force). Here, the changes in amplitude describing the diurnal fluctuations in physical activity were similar in magnitude, but changes in the time of peak levels, the acrophase, were minimal (***Supplementary Figure 7***).

The actigraphy device also allowed us to quantify wrist temperature ^20^. Traces clearly separated elevated nighttime wrist temperature readings from lower daytime ones with acrophases between 01:00 and 02:00 (***Supplementary Figure 8***). These findings remained stable over the course of the study (***Supplementary Table 4***).

From wearable ECG data, metrics of heart rate variability suggested elevated sympathetic nervous activity (SNS) (SNS Index MESOR preoperative 1.3[1.1] to 4.1[3.1] post-operative, *unadjusted p-value=*0.031) and loss in amplitude of successive heartbeats or RR intervals (74.7[52.6] to 39.1[23.9], *unadjusted p-value=*0.031) (***Supplementary Figure 9***, ***Supplementary Table 5***). Interestingly, several patients showed shifts in the acrophase of their mean RR intervals ranging between 2.3-3.1h for phase-advances (*unadjusted p-value*<0.003) and 3.8-8.8h for phase delays (*unadjusted p-value*<0.01) which can be conceptualized as patients experiencing acute jet lag while hospitalized (***Supplementary Figure 10***).

The metabolic rate estimated from ECG traces was elevated post-operatively by 122% compared to the preoperative baseline (means of median [IQR] energy kcal/min 1.8[2.5] to 4.0[2.9], ***Supplementary Figure 11***), a metabolic alteration common in patients after cardiac surgery due to the surgical trauma and its inflammatory response ^21^.

Vital signs measured continuously via an in-ear wearable device showed MESORs within physiological range for heart rate (mean±SD 74.0±13.4 bpm) and for tympanic temperature (36.7±3.2°C), but neither for respiratory rate nor oxygen saturation possibly caused by suboptimal fitting of the device (***Supplementary Figure 12***, ***Supplementary Table 6***). Per chart review oxygen saturation levels reached mean±SD 97.8±1.8% (***Supplementary Table 7***).

### Time-Specific Signals Emerge as Candidate Disruptors of Biological Rhythms

Based on door-mounted sensor data, patients received a median [IQR] number of 1[2] visit per hour between 01:00-05:00. Beginning at 07:00, traffic increased from 4[3.25] visits per hour to peak levels of 10[8.5] visits per hour at 11:00. In the afternoon and early evening hours, the frequency of visits hovered between 4-6 per hour [IQR 5-6], decreasing to 2 [IQR 1-2] visits per hour between 22:00 and 24:00 (***Supplementary Figure 13***). These data agreed well with observer-based assessments of room traffic (R^2^=0.62, ***Supplementary Figure 13***).

Patients were exposed in the patient room to sound levels reaching on average 51.9±3.3 dBA during daytime (06:00-22:00) and 48.3±4.2 dBA during nighttime (means calculated from hourly medians and SDs from hourly IQRs). Maximum median sound levels of 54.4 (3.7) dBA were reached between 11:00 and noon, while minimum median sound levels of 46.6 (4.2) dBA were evident between midnight and 01:00 (***Figure 1a***). These sound levels exceeded the recommended threshold of 45 dBA proposed for the hospital environment ^22^. Our observer-based reporting of alarm events did not correlate with patient room sound levels (R^2^=0.02) likely due to logging alarm sounds from both the patient room and the hallway care team workstation suggesting that hallway sounds do not affect patients.

**Figure 1.**
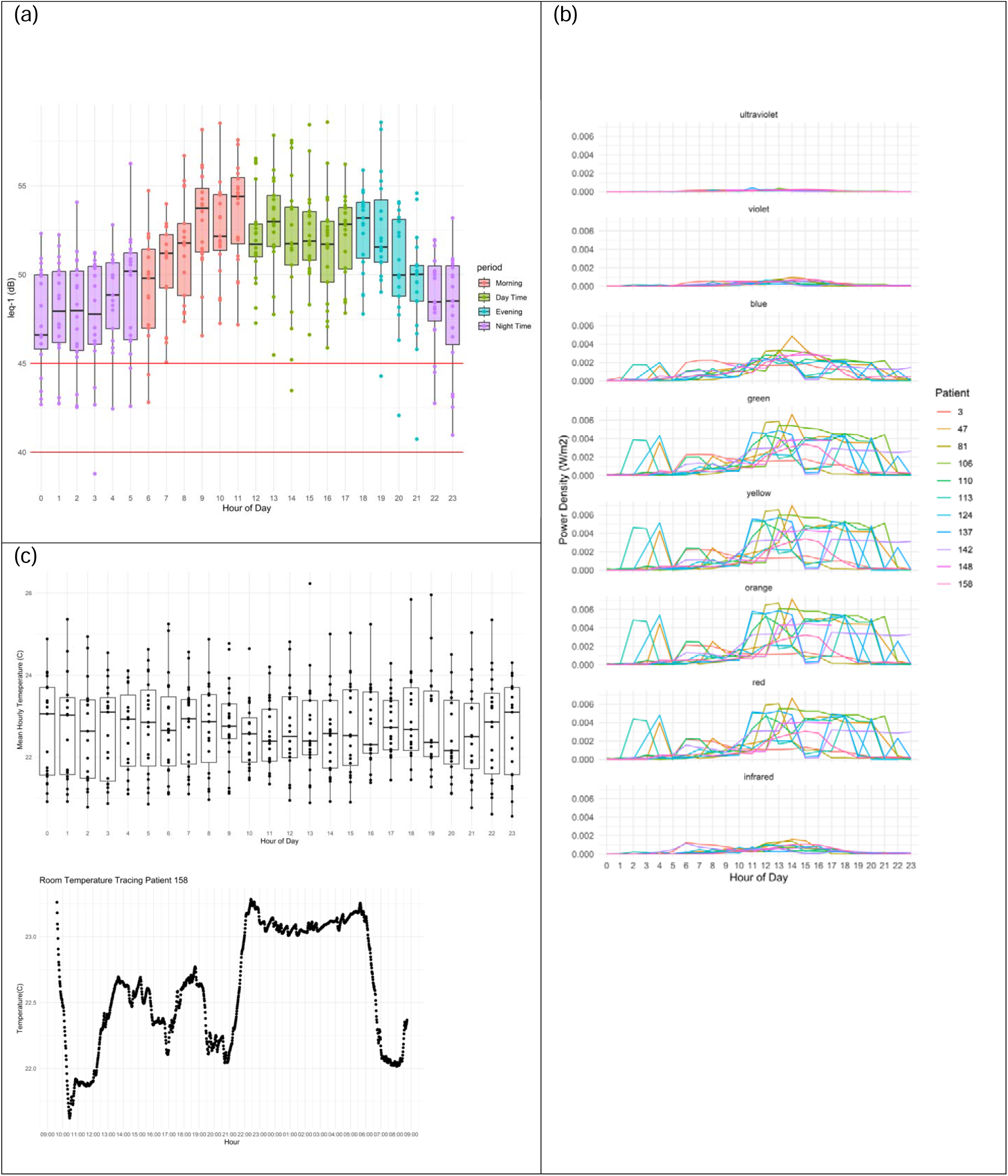
Patient Room Sound, Light & Temperature Levels. (a) Class 1 sound level meters were used as most accurate instruments to assess ambient sound exposure in the patient room. The red reference lines indicate thresholds at <45 dBA defined by the EPA specifically for hospital environments (Average equivalent sound level over 24-hr period, L_DayNight_ <45 dBA; noise during the nighttime hours 22:00-07:00 is penalized by adding 10 dBA to account for heightened noise sensitivity), and at <40db defined by the WHO for noise exposure during the night hours (Average outdoor nocturnal noise levels of LA_eq,outside_<40 dBA a long-term goal for the prevention of noise-induced health effects ^31^). The unit on the ordinate is the equivalent A-weighted sound level over a given time interval. Each point is the hourly mean of a patient at that hour of day from 2 days of monitoring (i.e. 2 points per patient at every hour). (b) A spectrometer device quantified the power density for each wavelength. Exposure to each spectral density (y-axis) is indicated for each patient by individual colors listed in the legend on the right and plotted over the 24 hours (x-axis) starting at “0” for midnight (c) Diurnal traces of ambient patient room temperature binned for each hour across the cohort (top) and plotted over the 24 hours (x-axis) starting at “0” for midnight with a single room-specific trace (bottom) where data collection started at around 10:00. Note the upswing at around 22:00 with highest room temperatures until the downswing at around 06:00.

The maximum brightness levels measured by a luxmeter as total intensity of visible light showed a typical diurnal distribution reaching 89.9±87.7 lux during daytime hours (calculated as mean±SD of hourly medians (IQRs)) which dropped to 3.7±9.8 lux at nighttime hours (defined as 22:00 to 06:00) (***Supplementary Figure 14***).

The light spectra in the patient rooms closely approximated natural sunlight during day hours (color rendering index [CRI] of 91.8±6.9 as the mean of hourly medians and IQRs where a CRI of 100 resembles natural sunlight). During night hours (22:00-06:00) the CRI dropped to 65.6±47. This pattern was reflected in the melanopic light exposure of 52.8±78.5 lux during daytime compared to 0.9±3.4 lux during night hours (melanopic light refers to specific wavelengths of light that activate melanopsin in the eye). Maximum melanopic light exposure of 115±75 lux occurred at 13:00 (***Supplementary Figure 15***). Correspondingly, mean blue light spectral power density of 25.5e^-4^±5.5e^-4^ W/m^2^ (Watt per square meter) was the highest measured at the same time of day. The daytime mean of blue light was 14.2e^-4^±7.3e^-4^ W/m^2^ versus 2.4e^-4^±3.9e^-4^ W/m^2^ at night. This pattern was similar for the green, yellow, orange, and red light spectral power densities. Notably, several patients were exposed to light in the early morning hours between 01:00-05:00 reaching similar spectral power densities as measured during peak daytime hours (***Figure 1b***).

Ambient temperature readings in the patient rooms did not display diurnal variability across the cohort (***Figure 1c top***). Mean daytime and nighttime levels were overall similar at 22.6±1.5 °C and 22.9±2.0 °C, respectively. A few patient room-specific traces illustrate the case, however, that ambient temperature showed diurnal variation with highest readings at night (***Figure 1c bottom***).

Finally, the room occupancy sensor indicated higher traffic in the private bathroom during the day compared to night (2.4±1.0 % versus 0.7±0.6 % bathroom-use-time normalized to device recording time, ***Supplementary Figure 16***) though these data cannot be associated to the patient specifically due to anonymous remote sensing.

Taken together, our assessments of the patients’ hospital environment suggest that room traffic, light and sound exposure and room temperature during night hours need to be considered as potential disruptors of behavioral, physiological and sleep-wake rhythms.

### Transient Postoperative Disruption of Behavioral & Physiological Biorhythms

To explore how the structure of behavioral biorhythms was affected by hospitalization, we used two common metrics to quantify the rhythm disturbances by calculating intra-daily variability and the robustness of rhythms by inter-daily stability ^23^. Physical activity calculated as vector magnitude in 30-sec epochs from wrist actigraphy trended to show higher rhythm disturbances with a median [IQR] for intra-daily variability of 1.8[0.3] in-hospital compared to baseline 0.8[0.4], *unadjusted p-value*=0.037. Rhythms were less robust for the in-hospital median [IQR] inter-daily stability of 0.5[0.3] compared to baseline median [IQR] 0.7[0.2], *unadjusted p-value*=0.063. After discharge, both activity disturbances and robustness of activity rhythms rebounded towards preoperative baseline levels (***Supplementary Figure 17***).

We furthermore used the wrist actigraph data to identify episodes of sleep where every 30-sec epoch over 24 hours was labeled as either asleep (1) or awake (0). These data also allowed calculation of disturbances (intra-daily variability or IV) and robustness (inter-daily stability or IS) of sleep rhythms. Here, in-hospital sleep rhythms were more disturbed (0.8[0.2]) compared to preoperative baseline (IV of 0.4[0.1], *unadjusted p-value*=0.002). Sleep rhythms were less robust for the in-hospital condition (0.7[0.3] compared to preoperative baseline of 0.9[0.1], *unadjusted p-value*=0.0059, ***Supplementary Figure 17***). While robustness of sleep rhythms after discharge rebounded towards preoperative baseline levels, sleep rhythm disturbances post discharge stayed elevated at an IV of 0.9[0.3] compared to the preoperative baseline condition of 0.4[0.1], *unadjusted p-value*=0.0098.

Next, we used outputs from the cosinor fits of accelerometer data from wrist-worn devices to correlate MESOR and amplitude of the vector magnitudes as proxy for physical activity. We specified MESOR as the independent variable since only high levels enable larger absolute daily swings in the amplitude. Consequently, pre-operatively a large proportion of the variation in amplitude was predictable from the MESOR values (R^2^=0.89). In-hospital, this relationship deteriorated (R^2^=0.28) but showed a rebound post discharge (R^2^=0.65) (***Supplementary Figure 18 top***). Notably, the MESOR-amplitude relationship of physical activity of the present cohort (R^2^=0.89) at baseline resembled closely the one we measured with a similar wrist-worn device in apparent healthy young (R^2^=0.79) and old volunteers (R^2^=0.85) (n=10 per group, manuscript under review) (***Supplementary Figure 18 bottom***).

Finally, we explored disturbances (inter-daily stability) and robustness (intra-daily variability) of wrist temperature rhythms to find a drop in robustness from baseline 0.8[0.3] to in-hospital 0.5[0.3], *unadjusted p-value*=0.049, while the degree of rhythm disturbances of wrist temperature remained similar (*unadjusted p-value*=0.5, ***Supplementary Figure 19***).

### Transient Postoperative Disruption of Sleep

Polysomnographic evaluation of wearable PSGs deployed during the hospital stay indicated a deficiency of Stage 3 and rapid eye-movement sleep as evident in the hypnogram from a selected patient (***Figure 2a***). The ORP derived from the electroencephalogram power spectrum analysis ^15,16^ provided further insight by reporting sleep depth as a single index ranging from zero (very deep sleep) to 2.5 (full wakefulness). This index showed the right skewed ORP stages, comprised of increased levels of full wakefulness, drowsy awake and transitional sleep during night time compared to published healthy ORP sleep architecture ^16^ (***Figure 2b***). Across the cohort, ORP staging revealed that a median [IRQ] of 21.0[30.0]% of time was spent in full wakefulness during the night compared to the recommended 5% or less of time as published ^16^. Similarly high percentages were spent drowsy awake (16.6[6.0]%) and in transitional sleep (23.6[10.2]%) compared to published normative durations of 5% and 10% ^16^, respectively. Time spent at night in deep sleep (6.5[6.4]%) and average sleep (7.2[8.4]%) was considerably less than the close to 25% of time expected ^16^. Strikingly, the statistical comparison of ORP stages between night and day times did not suggest differences (*unadjusted p-values*>0.05) (***Figure 2c***).

**Figure 2.**
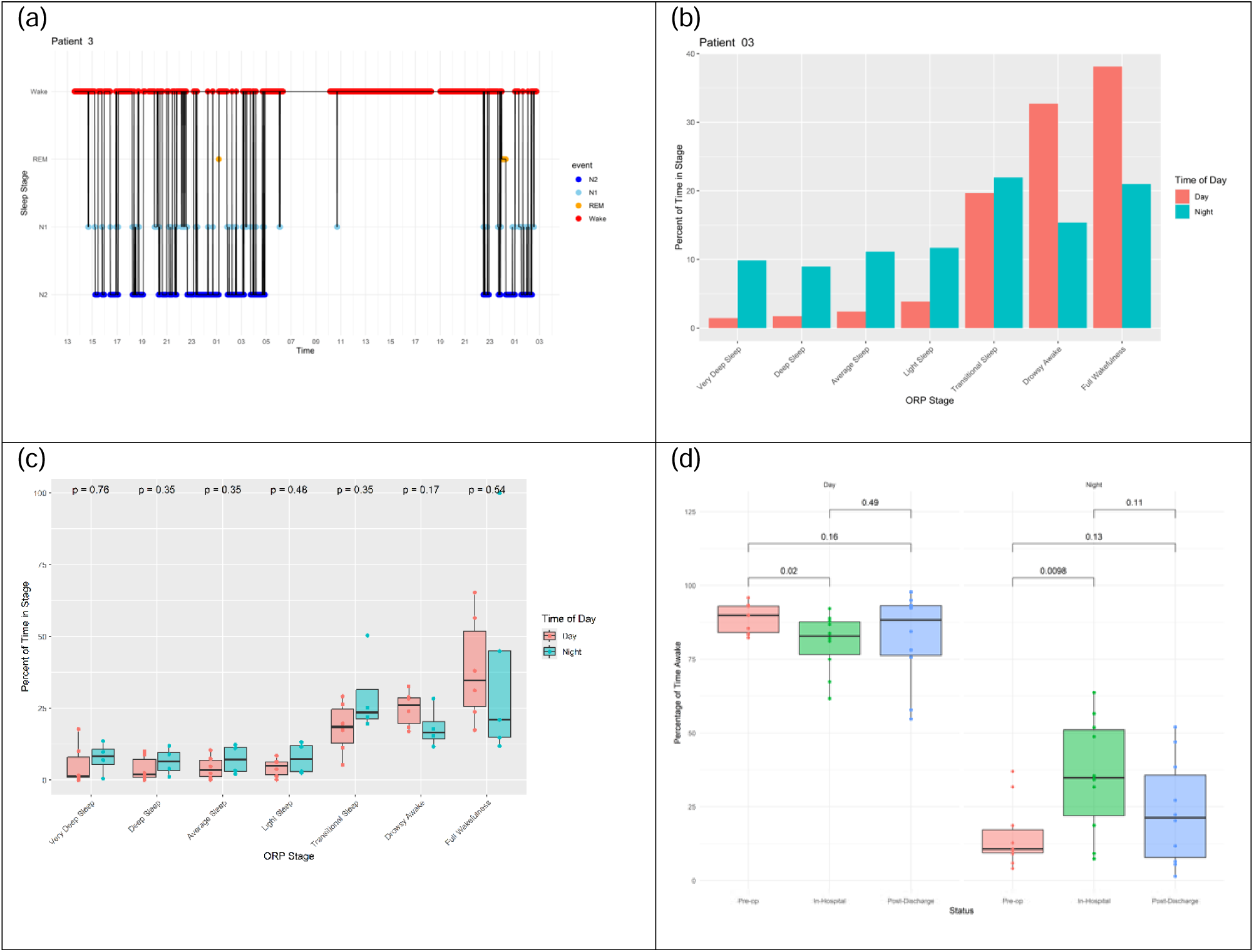
Disrupted Sleep-Wake Rhythms. PSG traces were collected through a wearable device which provided hypnograms shown in (a) for an individual patient along with the odds ratio product (ORP) time spent in sleep stages for that same patient in (b), as well as the ORP distribution across the cohort in (c). P-values resulting from unpaired Wilcoxon tests. Sleep/wake estimates derived from wrist accelerometry are visualized in (d) with percent wake time during the day (left) and night (right) for the pre-operative, in-hospital and post-discharge conditions. P-values resulting from paired Wilcoxon tests.

To address the limitation that PSG recordings were only available in a subset of patients (6 out of 13), we derived sleep-wake estimates from wrist actigraphy using published machine learning algorithms ^17^. Validating these estimates within patients with their PSG data showed good agreement (AUROC of 0.81, 95% CI: 0.81-0.82). The percentage of time spent awake during the day decreased from 89.8±9.0% at the preoperative baseline to 82.9±11.1% in-hospital (*unadjusted p-value=*0.02 Wilcoxon rank sum test). Conversely, time spent awake during the night hours increased from 10.7±7.9% preoperatively to 34.8±29.1% in the hospital (*unadjusted p-value=*0.0098 Wilcoxon rank sum test). Both trends rebounded after discharge (***Figure 2d***).

### Transient Post-Operative Decline of Cognitive Function

The PVT, administered through a mobile phone app, produced preoperative reaction times of 431±104 ms in the morning, 412±132 ms at midday and 467±69 ms in the evening, suggesting that alertness was highest at midday. Alertness was overall lower in the hospital environment, where reaction times were slower by 19.2%, 34.9% and 33.3%, respectively (corresponding to absolute increases of 514±107 ms in the morning, 555±73 ms at midday and 623±130 ms in the evening). Reaction times started to rebound once patients were at home. A substantial time-specific difference in reaction time was noticeable for in-hospital morning versus evening values (*unadjusted p-value=*0.011 unpaired Wilcoxon rank sum test) (***Figure 3a*).**

**Figure 3.**
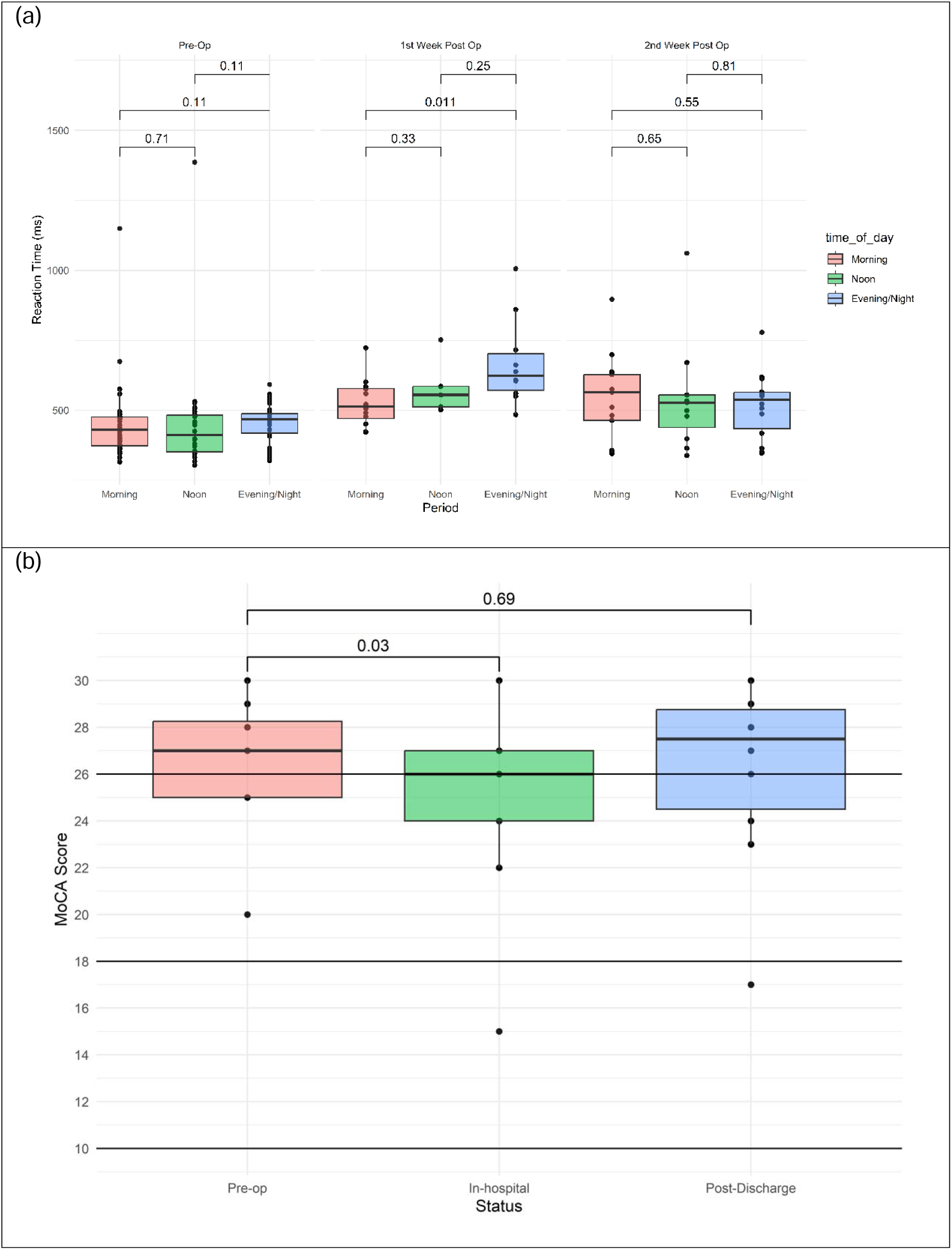
Post-operative Decline in Cognitive Function. (a) Reaction times from the Psychomotor Vigilance Test (PVT) administered through a phone mobile app assessed three times per day (morning, noon and evening/night) and (b) scoring from the Montreal Cognitive Assessment (MoCA) during the pre-operative, in-hospital and post-discharge conditions. P-values in (a) result from paired Wilcoxon tests. P-values in (b) result from multivariate mixed effects linear regression. A score of 26 points represents the threshold level for a normal score, a score of 18 points is the threshold for moderate impairment and a score of 10 points is the threshold for severe impairment.

To screen for cognitive impairment in our study participants, we deployed the Montreal Cognitive Assessment (MoCA, paper version) screening test. We found that the MoCA score decreased by two points from a mean±SD of 26.8±2.8 points preoperatively to 24.7±3.9 points in-hospital (the mean delta change was -2.3 points between baseline and in-hospital). Cognitive function recovered across the cohort after discharge back to a score of 26.2±4.0 points (***Figure 3b).*** Notably, about 31% (4/13) of patients showed in-hospital declines in their MoCA score exceeding 1SD (calculated from the baseline MoCA assessments) categorized as mild impairment (***Supplementary Figure 20***). This phenotype is consistent with what has been described in the literature as postoperative cognitive dysfunction (POCD) ^24^. In the SDU, delirium was not detected using the confusion assessment method (3D-CAM).

## Discussion

This pilot study established the feasibility of a comprehensive time-specific characterization of behavioral, physiological rhythms and health outcomes of elective cardiac surgery patients in the hospital environment. The main findings highlight how patients transitioning from their preoperative home environment into the hospital environment post-surgery experienced disrupted behavioral, physiological and sleep-wake rhythms paired with a transient decline in cognitive function. We identified room traffic, sound and light exposure as well as room temperature, especially during nighttime, as modifiable sources of potential circadian disruption. Examining exposure-outcome relationships suggests an association between higher room traffic, sound, light and temperature levels at nighttime and higher percentages of time awake during night hours. As shown in ***Figure 4*** several patients with a ∼1SD higher exposure showed large increases in wakefulness during the in-hospital night compared to preoperative nights. The acquisition of these data will inform the design of a randomized trial to assess the impact of chrono-mitigation in the hospital on clinical outcomes.

**Figure 4.**
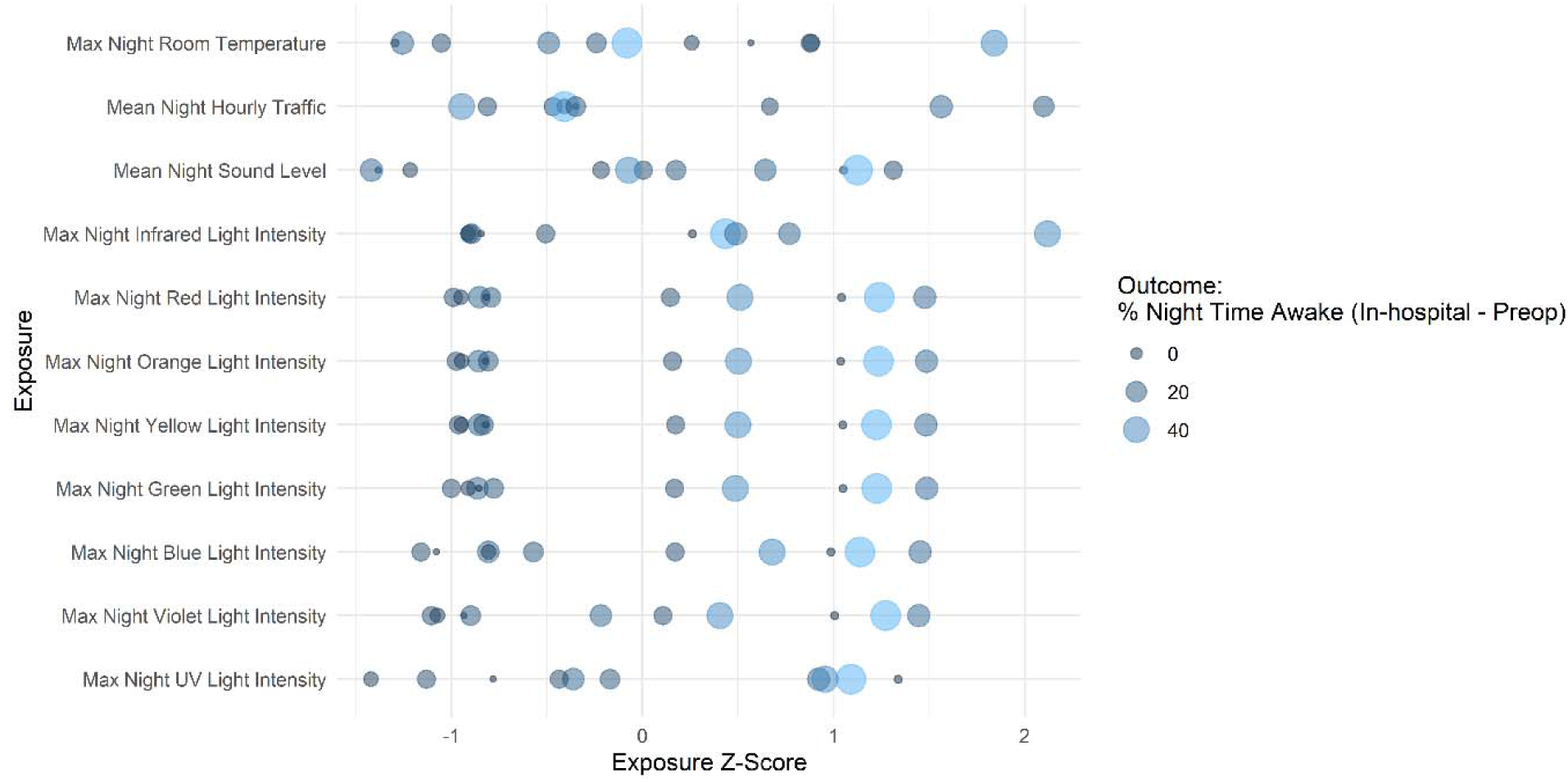
Exposure-Outcome Relationships. Dot plot of patients’ exposure to ambient room temperature levels, room traffic, sound and light levels at nighttime, normalized by z-scoring, versus patient outcome defined as percentages of time awake during night hours for the change in-hospital compared to pre-operative baseline. Percentage of time spent awake was obtained from wrist accelerometry. Dot sizes and colors are both proportional to effect sizes of the outcome variable. Despite low sample size and considerable variability, a trend emerged where larger dots indicating more time spent awake in-hospital were associated with higher z-scored exposure to room traffic, sound levels and light intensities at night.

A particular strength of our study is that it demonstrates the feasibility of in-depth characterization of diurnal phenotypes by using redundant but orthogonal approaches. Eleven wearables, nearables and mobile phone apps collected longitudinal data at high resolution over days up to a week in these subjects. Different device platforms were used for several clinically relevant study endpoints. For example, sleep-wake rhythms were assessed by both PSG and wrist actigraphy, the latter parsed by machine learning algorithms to deliver sleep/wake estimates. Different biological systems regulated by circadian clock machinery were interrogated to quantify effects on physiological rhythms, such as heart rate variability and body temperature. Furthermore, we extended surveys with remote sensing technology to assess dimensions of cognitive function, for example, through the MoCA screen test paired with the PVT over the phone app.

Our data are consistent with the literature where it is available. Sound readings in hospital environments at peer institutions produced comparable levels at 52-54 dBA ^25^. As expected, reaction time in the PVT from our patients was much slower to those in airline pilots ^26^. PVT results in our patients followed a u-shaped pattern over the course of the day with slower reaction times in the morning, fastest at midday, and slower again in the evening. This diurnal phenotype has been described in keystroke dynamics assessed on the mobile phone’s keyboard and was most pronounced in elderly participants ^27^. Furthermore, this diurnal phenotype was evident in response times to noncognitive ecological momentary assessments (EMA) ^28^. Since the PVT was administered through a mobile app under real world conditions, settings were not standardized. Thus, PVT likely integrated location- and patient-specific factors such as distractions, motivation, mood and medication as well as reduced attention from sleep loss and desynchronized biological rhythms.

Somnuke P *et al.* ^24^ recently enrolled patients ≥ 60 years of age with neurological, cardiothoracic, colorectal, hepatobiliary, gynecological, urological, and orthopedic surgical procedures into a study with a similar design but larger sample size (n=279) where they administered MoCA screen test preoperatively to determine baseline cognitive status and repeated the screen between 5 and 9 days after surgery. The authors characterized postoperative cognitive dysfunction by a MoCA score reduction of 2 or more points from the preoperative score and found that this condition occurred in 24.3% of patients with a preoperative diagnosis of mild cognitive impairment but in 50% of patients with normal cognition prior to surgery. This study provides the framework to categorize the transient cognitive decline in our cohort as postoperative cognitive dysfunction and thus highlights the clinical relevance of our findings. While a mild transient decline in cognitive function after major surgery under general anesthesia medicated with opioids and sedatives may be perceived as expected and harmless, recent evidence suggests a risk associated with this specific phenotype. A prospective study of 167 older adults undergoing major noncardiac surgery found that patients with postoperative cognitive dysfunction were twice as likely to experience new impairments in instrumental activities of daily living compared to patients without postoperative cognitive dysfunction ^29^.

We designed this study with the understanding that we introduced three specific biases into the clinical protocol: First, we selected patients scheduled for elective cardiac surgery with the expectation that this type of surgical procedure followed by mandatory hospitalization in critical care followed by an acute care SDU skews this patient cohort towards detection of disruptions in their biological rhythms and health outcomes. Second, we chose to schedule study assessments to occur during the acute re-stabilization period in the SDU. We reasoned that the SDU environment is more easily modified in an interventional study than the immediate post-operative period in the ICU where patient vulnerability necessitates frequent care delivery. Third, we biased against detecting temporal disruption by conducting this study in a recently completed facility purposefully designed to enhance the patient care experience ^30^.

This study has several limitations. As a pilot study to generate preliminary data the sample size is small, so that it is unclear if our results are generalizable. Though we enrolled generally healthy patients with normal preoperative cognition and good postoperative recovery, the sample included a broad array of cardiac surgery procedures which likely introduced substantial noise into our measurements. This approach, however, reflects a real-world operating schedule which facilitates scalability for future studies. The *p*-values we report are not corrected for multiple testing and therefore only indicate trends to support interpretation of plots and summary statistics. The effects of medications on time-specific phenotypes cannot be determined in the present study due to the myriad possible interactions dependent on the active drug, its dose, metabolism, route of administration, and timing. Finally, a key objective was to assess the feasibility of collecting such data, directly and remotely in a postoperative environment. Upon arrival in the SDU, we scheduled deployment of wearable devices after all standard acute patient care had already been implemented. While all devices were deployed to most patients, several patients declined wearing the PSG device citing a concern of intrusiveness. During the 48-hour of device data collections in the SDU, the study team usually interacted with the study participant 2-3 times per 24-hour interval for surveys, to facilitate study compliance, and to exchange the PSG and in-ear vital sign devices (to comply with the need to recharge batteries). We noted during these visits that the PSG device often slid down on the patient’s forehead towards the glabella (3 out of 6 placed devices), which led in some cases to device removal by the patient when this happened during the night. Similarly, we noted that an in-ear vital sign device was dislodged from its proper position in the ear canal (1 out of 9 placed devices). These conditions introduced noise into our dataset and could cause time points to be missed.

In conclusion, we show pilot evidence of desynchronized behavioral, physiological and sleep-wake rhythms for patients undergoing elective cardiac surgery in the hospital environment accompanied with a transient decline of cognitive function. Patient room traffic, sound, light and room temperature emerge as modifiable circadian/diurnal disruptors. We will use this information to address the practicality and value of chronomitigatory strategies in a hospital environment.

## Supporting information

STROBE checklist

## Data Availability

The data used for the analysis are accessible at www.tbd.org.

## Code Availability

The code used for the analysis is accessible at www.tbd.org.

## Acknowledgments

We are especially indebted to the patient volunteers, their families, the patient care teams at the Hospital of the University of Pennsylvania Heart & Vascular ICU & the Pavilion 9 Center Cardiac Surgery Progressive Care. We extend our thanks to Michelle Feil, Ron Anafi, Mathias Basner, and Rebecca Trotta for helpful discussions. LaVenia Banas supported this study as clinical study coordinator. Our observers on the floor included Amarachi Mbadugha, Angela Zhu, Camille Quaye, Chelsea Chin, Dineth Karunamuni, Kamille Hernandez, Marbella Aguilar, Mary Londono-barrios, Mohammad Abrar, Neil Reddy, Olivia Ye, Rose Kim, Sandy Li, Shikhar Gupta and Tharunika Vuppalapatti Sekar. Funding was kindly provided by the Clinical and Translational Research Award (2U54TR001878) and the University of Pennsylvania Health System (UPHS). We are most grateful to Mr. Kevin Mahoney, CEO of UPHS who has provided sustained support for this study from the time of its conception. M.C. was supported by the National Institute of Nursing Research (R00 NR019862).

C.S. is the Robert L McNeil Jr. Fellow in Translational Medicine and Therapeutics. G.A.F. is the Robert L McNeil Jr. Professor in Translational Medicine and Therapeutics.

The concept for this study was presented by CS as a poster titled “Circadian Rhythm Disruption in the Hospital Critical Care Environment: Approaching Characterization in a Pilot Study” at the Society for Research on Biological Rhythms’ 2024 Biennial Meeting, held May 18-22 in San Juan, Puerto Rico. Amarachi Mbadugha successfully developed from the data an independent study project leading to her honors thesis on “Quantifying Circadian Program Disruptions in ICU Patients: A Pilot Study of Heart Rate Variability in the Hospital of the University of Pennsylvania” in the Department of Biology, University of Pennsylvania and sponsored by CS and Dr. Philip Rea, Professor of Biology in the Department of Biology in the School of Arts and Sciences, and Rebecka and Arie Belldegrun Distinguished Director of the Life Sciences & Management Program.

## Author Contributions

CS and GAF designed the study and obtained ethics approval. NEJ, MG, PA, NDD, and MAA recruited study participants with support from CS. NEJ, CS and GAF analyzed the data and interpreted the results with support from NFL, TGB and AMR. CS and GAF wrote the manuscript. NEJ and CS prepared figures and tables. All authors reviewed the manuscript and provided feedback which was the basis for several rounds of revisions by CS, NEJ and GAF prior to submission.

## Competing Interests

The authors declare no competing financial interests.

## Supplementary Materials

### Supplementary Figures

**Supplementary Figure 1.**
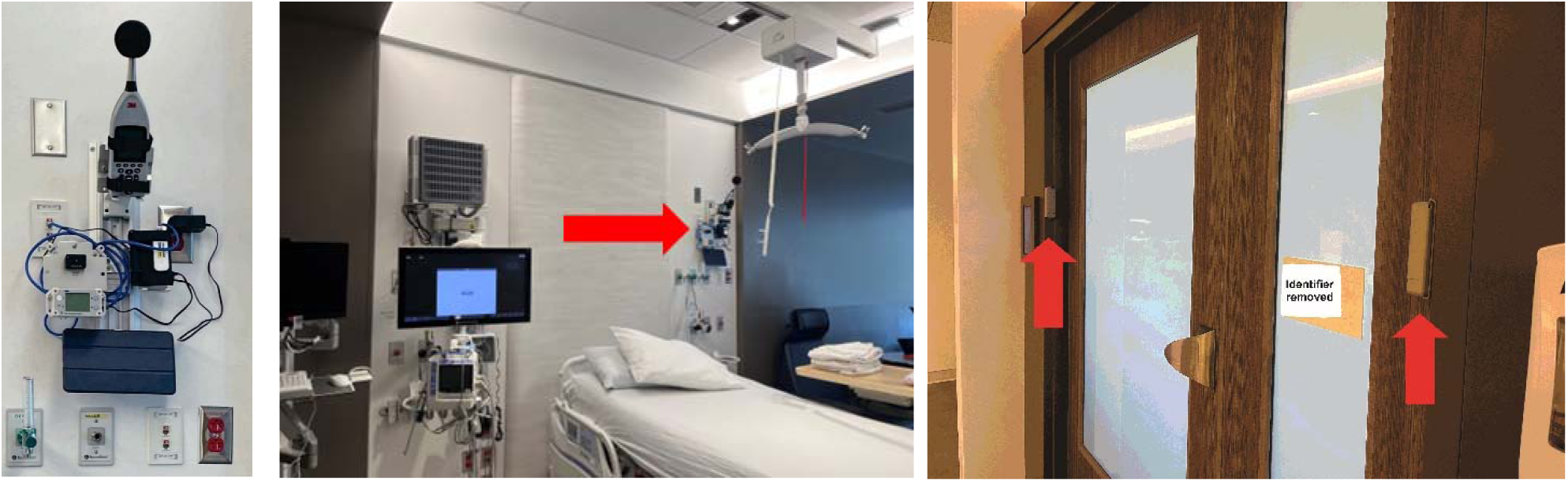
Chronobiome Weather Station. Standardized sensor setup in the patient room mounted on hospital-grade channel mounts (left) and patient room door (right).

**Supplementary Figure 2.**
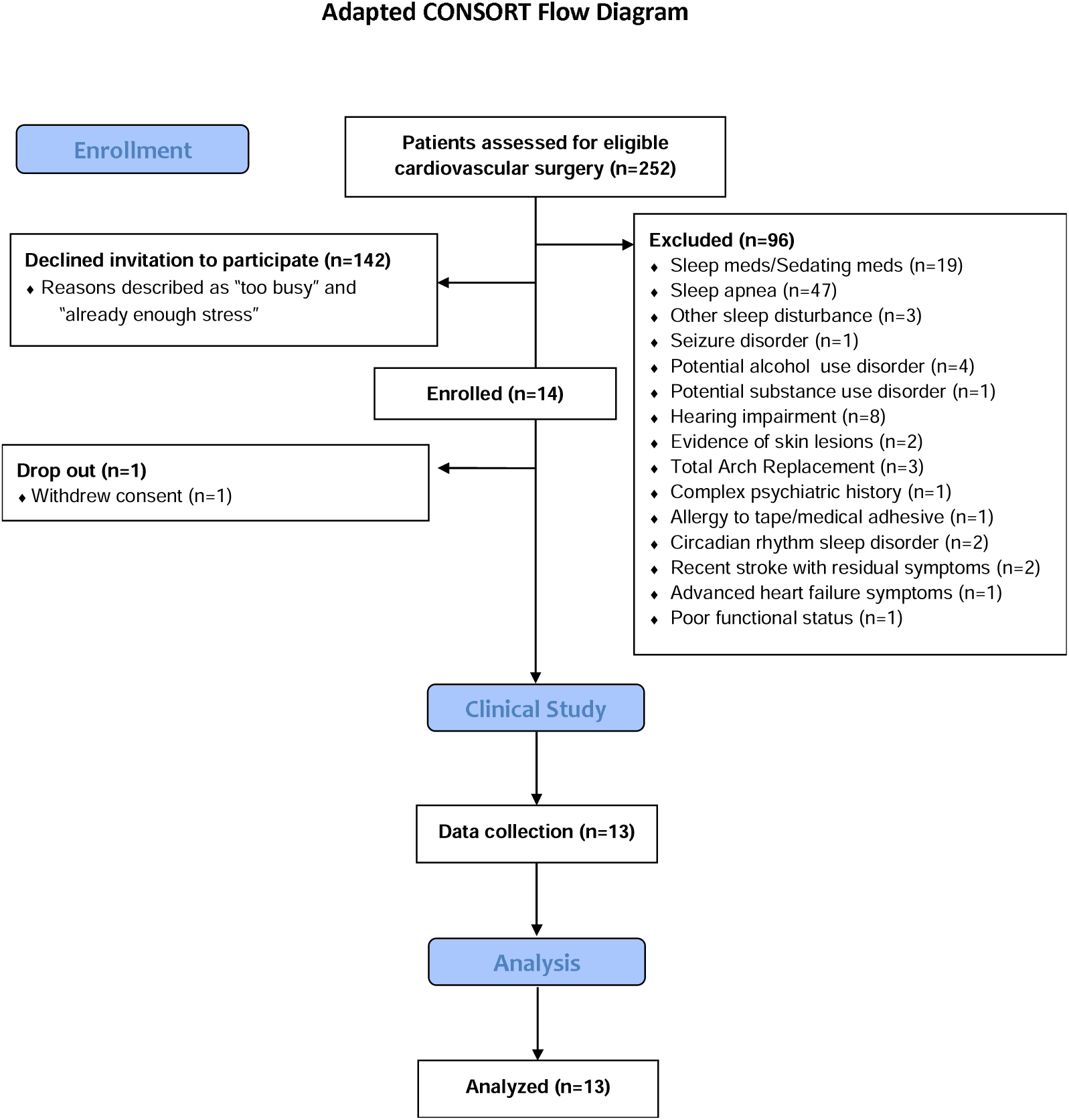
Adapted CONSORT Flow Diagram.

**Supplementary Figure 3.**
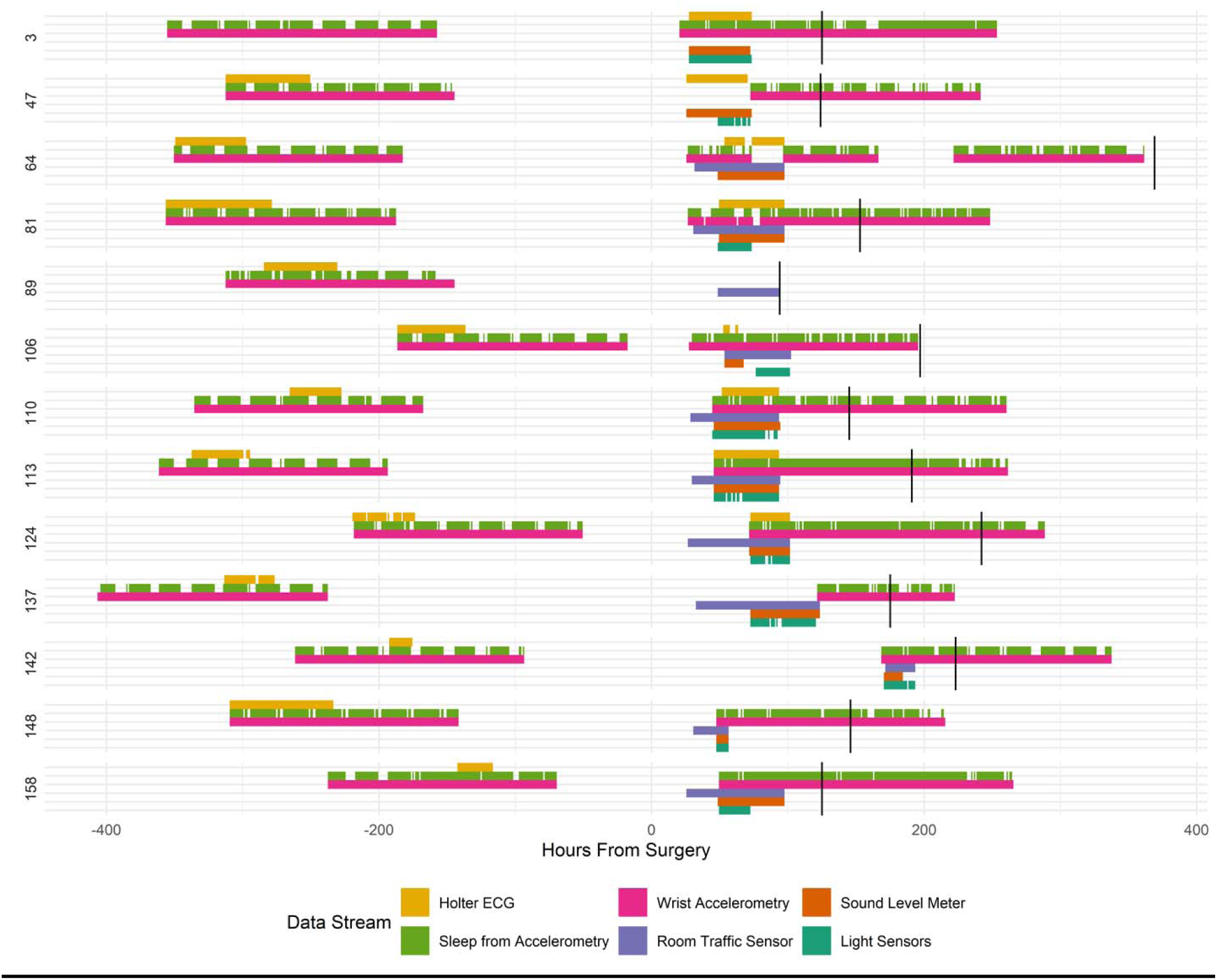
Data Coverage. Representative data collections for selected data streams from patient room sensors and patient wearables (legend) visualized per patient (right y-axis) in reference to time of surgery (zero on the x-axis). Each vertical tick (tile) represents any data present in that one hour. The vertical bar post-surgery indicates the time of discharge.

**Supplementary Figure 4.**
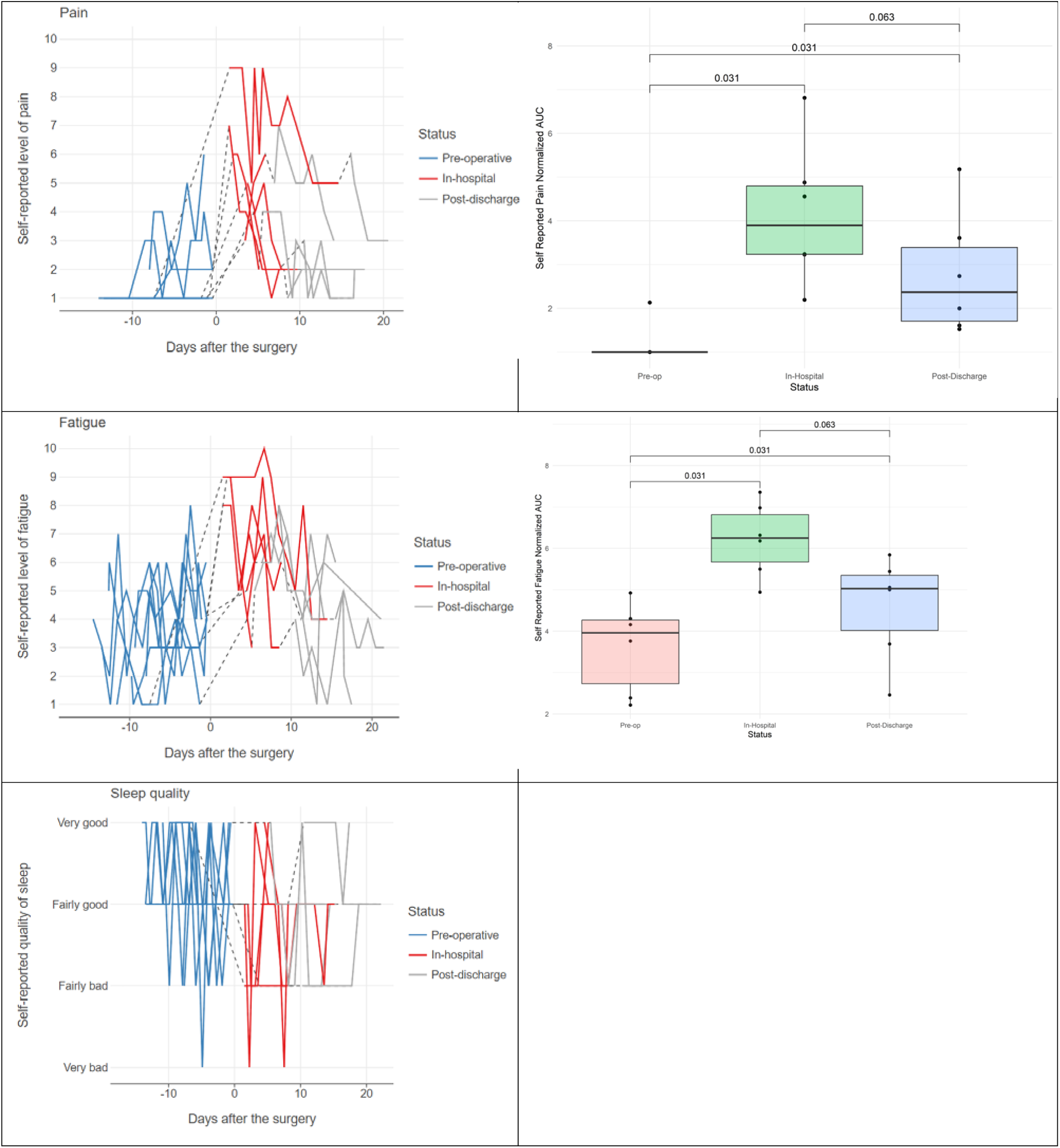
Self-Reported Pain, Fatigue, Sleep Quality. Self-reported levels of pain and fatigue administered daily through a mobile phone app. (Left) Levels of self-reported pain, fatigue and sleep quality over time pre-operatively, in-hospital and post-discharge where zero on the x-axis denotes the day of surgery. (Right) Boxplots of the normalized area-under-the-curve (AUC) of self-reported pain and fatigue categorized into pre-operatively, in-hospital and post-discharge. P-values result from paired Wilcoxon tests.

**Supplementary Figure 5.**
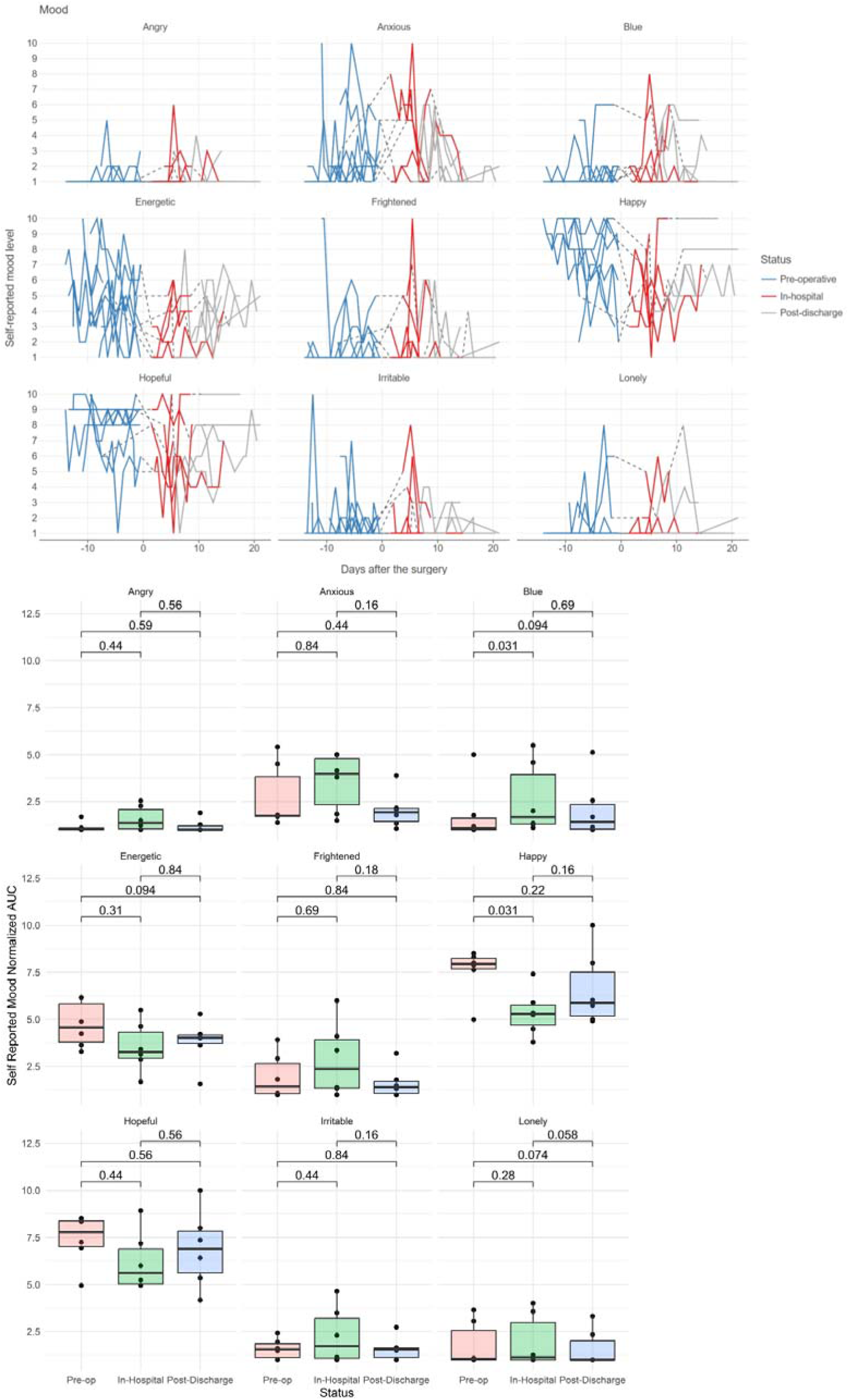
Emotional Valence. Self-reported levels of nine dimensions of mood administered daily through a mobile phone app. (Top) Levels of self-reported mood over time pre-operatively, in-hospital and post-discharge where zero on the x-axis denotes the day of surgery. (Bottom) Boxplots of the normalized area-under-the-curve (AUC) of self-reported mood categorized into pre-operatively, in-hospital and post-discharge. P. values result from paired Wilcoxon tests.

**Supplementary Figure 6.**
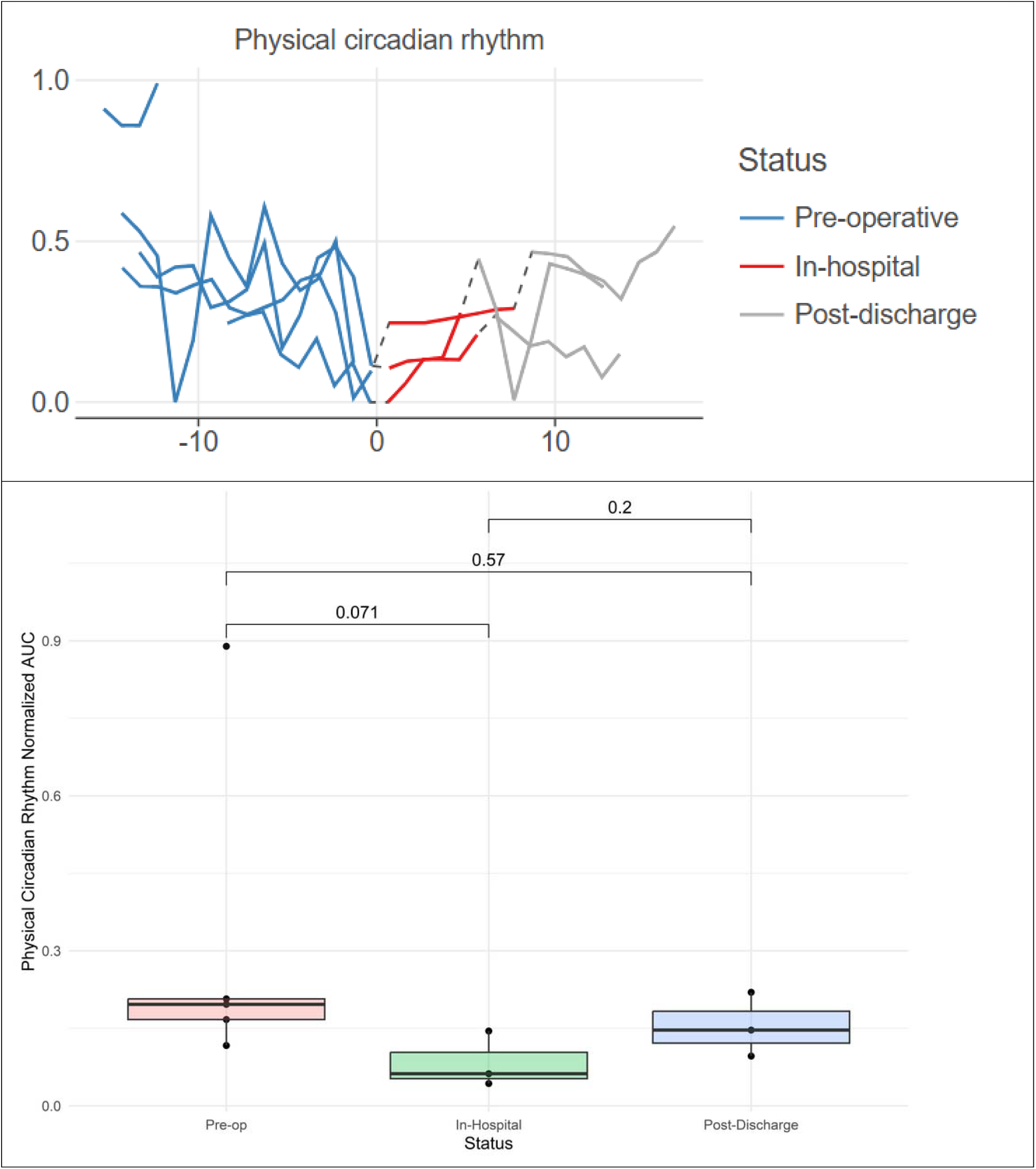
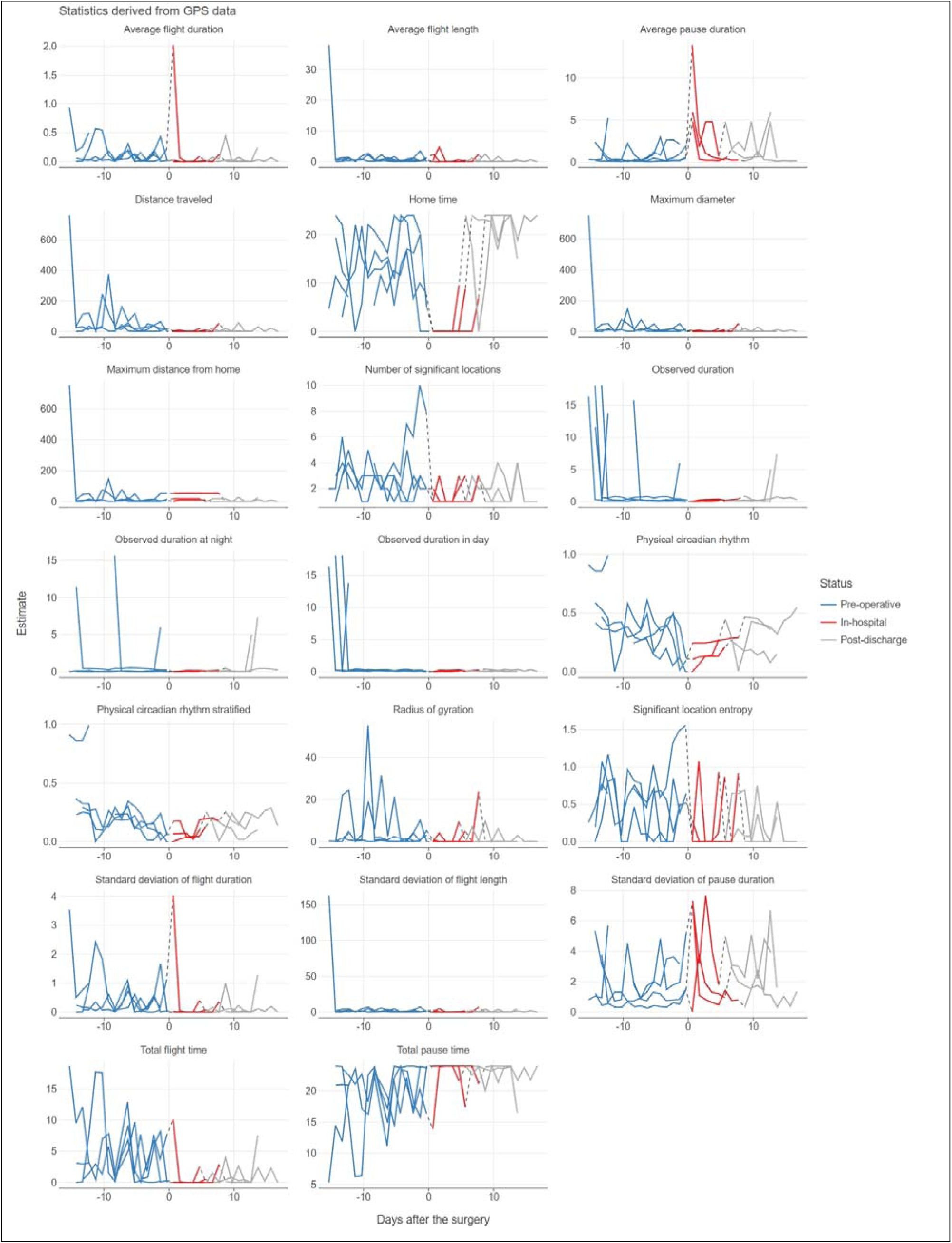
Disrupted Daily Routine & other GPS-based Outputs. (Top) Mobile phone-derived geolocation data transformed into the parameter “Physical Circadian Rhythm” which is a continuous measurement of routine where 0 on the y-axis indicates a complete break from routine and where 1 signifies the exact same routine from day to day. (Left) Levels of “Physical Circadian Rhythm” over time pre- and post-operatively where zero on the x-axis denotes the day of surgery. (Center) Boxplots of the normalized area-under-the-curve (AUC) of “Physical Circadian Rhythm” categorized into pre-operatively, in-hospital and post-discharge. (Bottom) Other GPS-based outcome variables collected through the Beiwe smartphone app ^12^. P-values result from paired Wilcoxon tests.

**Supplementary Figure 7.**
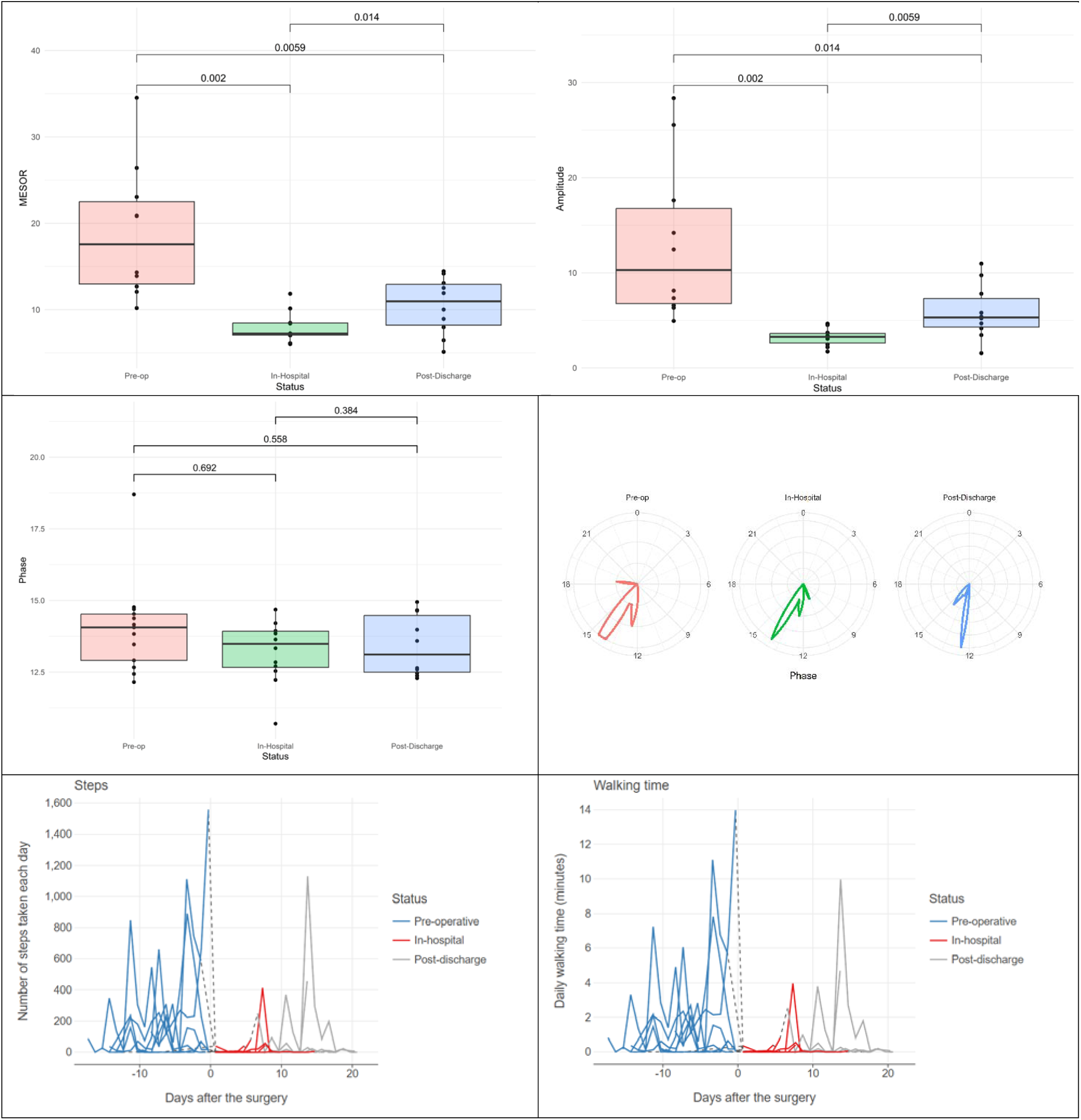
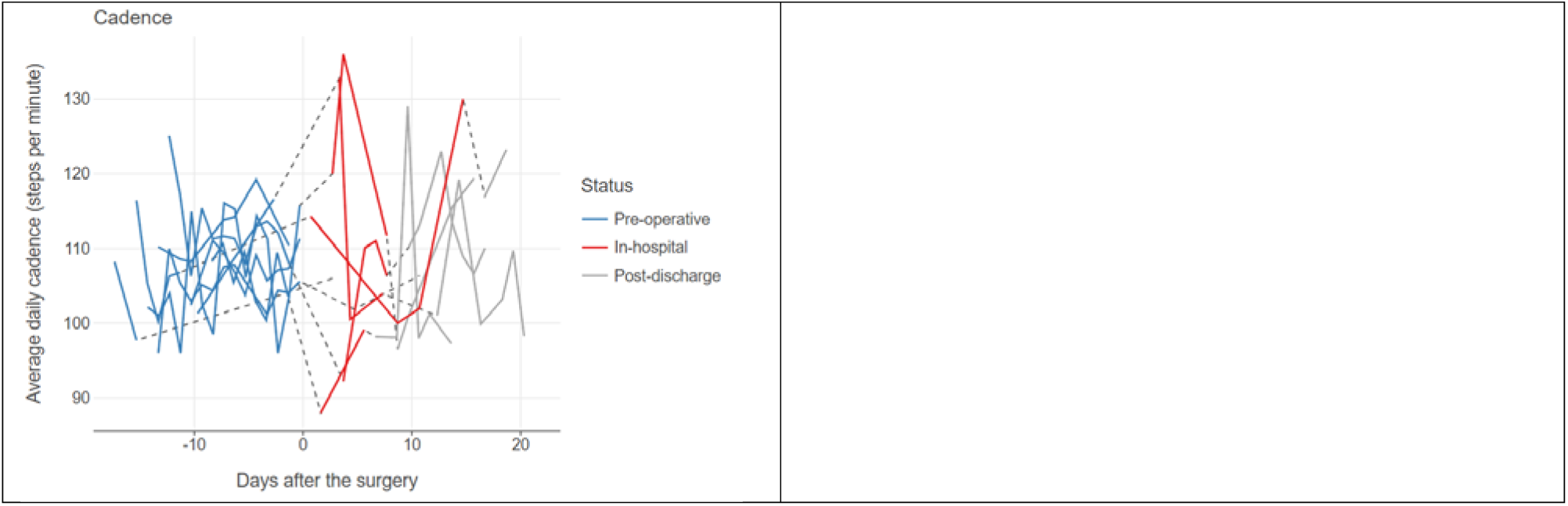
Physical Activity. Cosinor fits of accelerometer data from wrist-worn activity trackers provided (top) MESOR, amplitude, and (center) acrophase for the pre-operative, in-hospital and post-discharge conditions. (Bottom) Beiwe smartphone app-based step counts, walking time and cadence. P-values result from paired Wilcoxon tests.

**Supplementary Figure 8.**
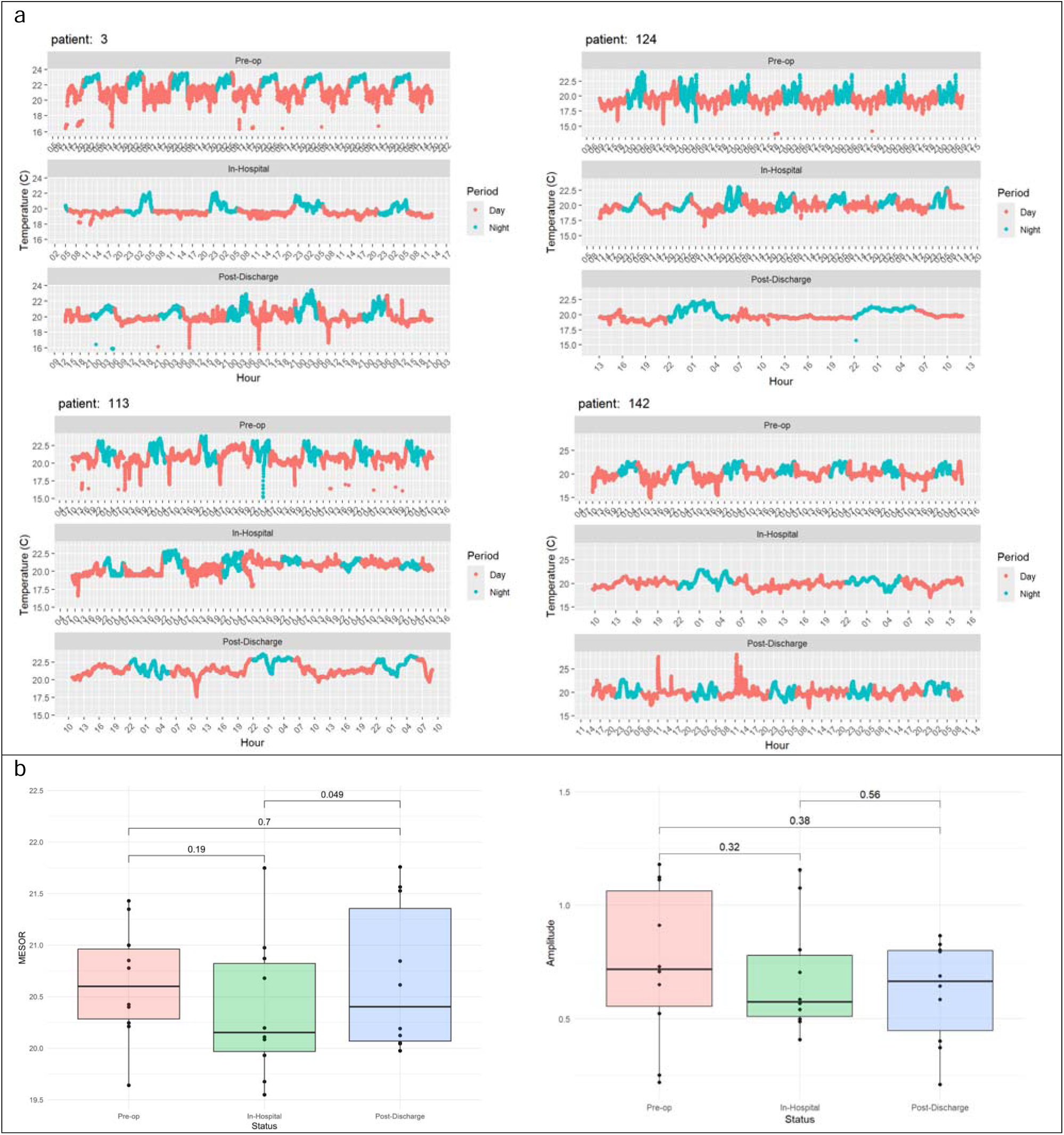
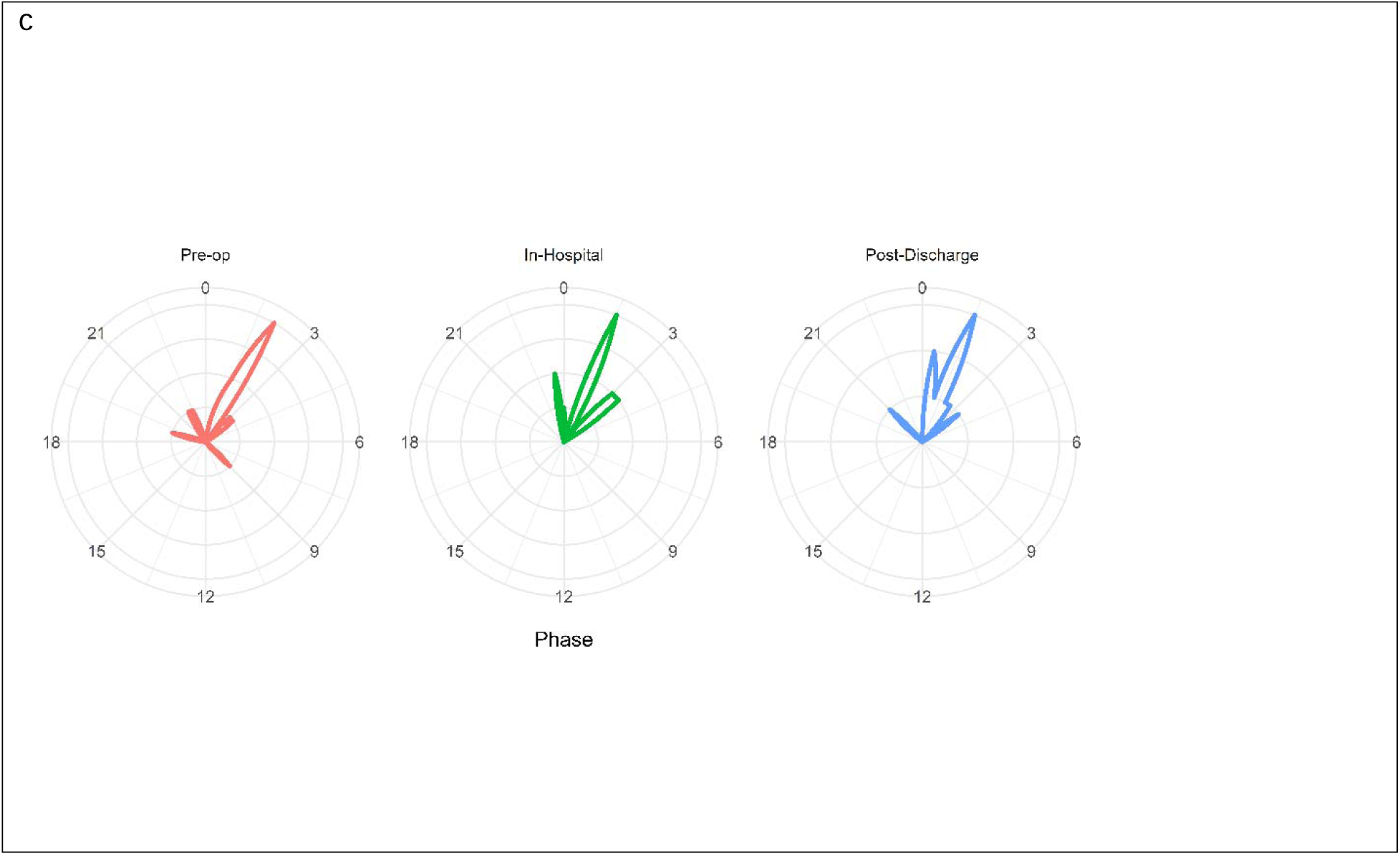
Wrist Body Temperature. Cosinor fits of wrist temperature data extracted from wrist-worn activity trackers provided MESOR, amplitude, and acrophase for the pre-operative, in-hospital and post-discharge conditions, colored for day (0600-22:00) and night times for individual study participants (a) and cohort (b, c). Note that the traces of the four study participants representative of the diurnal variability observed in wrist temperature data were selected based on the low degree of data missingness. P-values result from paired Wilcoxon tests

**Supplementary Figure 9.**
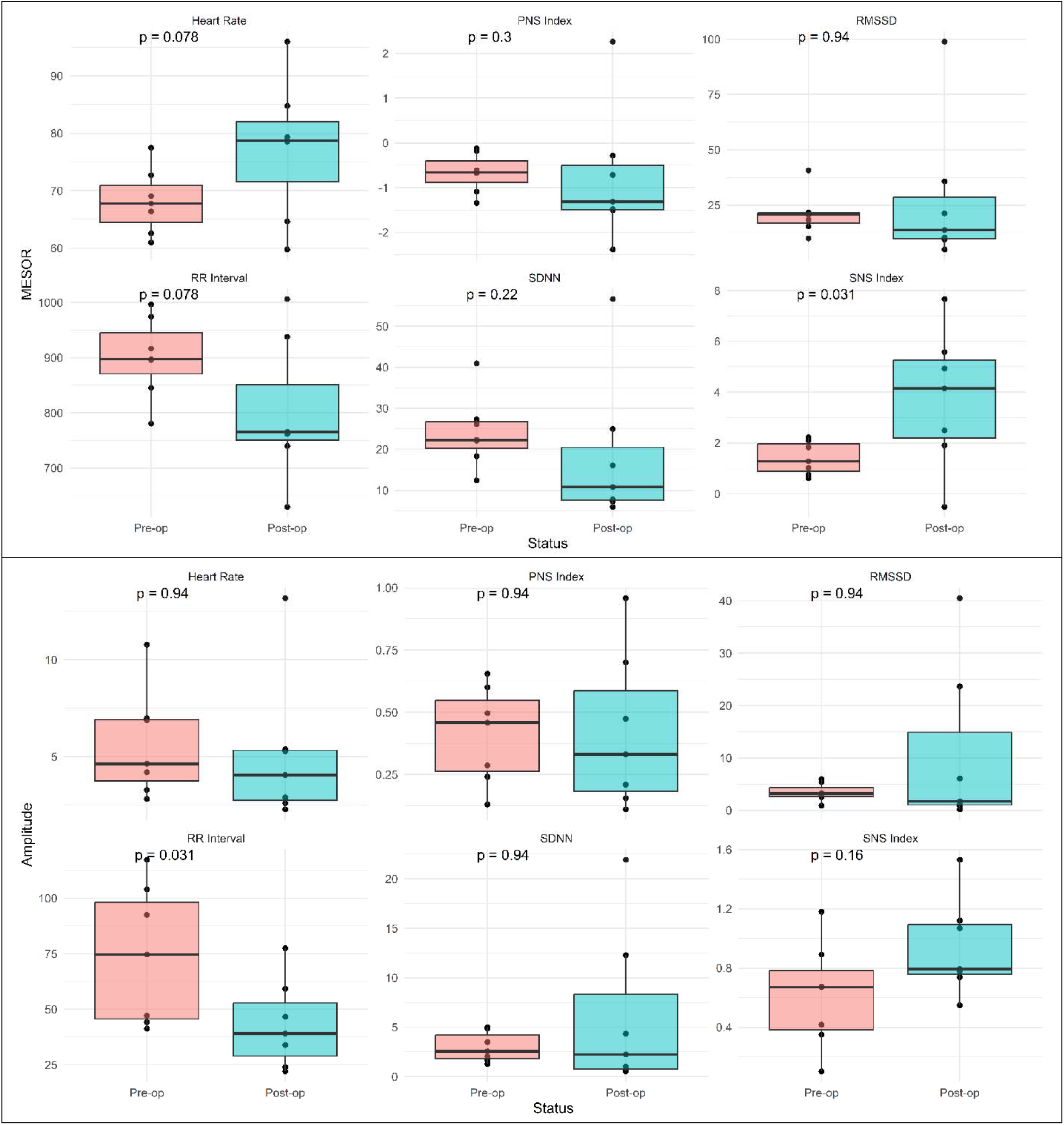
Cardiovascular System. Cosinor fits of ECG data collected with a chest-worn wearable device provided MESOR, amplitude, and acrophase of heart rate, heart rate variability (RR interval, SDNN, and RMSSD) as well as autonomic function (SNS and PNS index) for the pre-operative and in-hospital conditions. P-values result from paired Wilcoxon tests

**Supplementary Figure 10.**
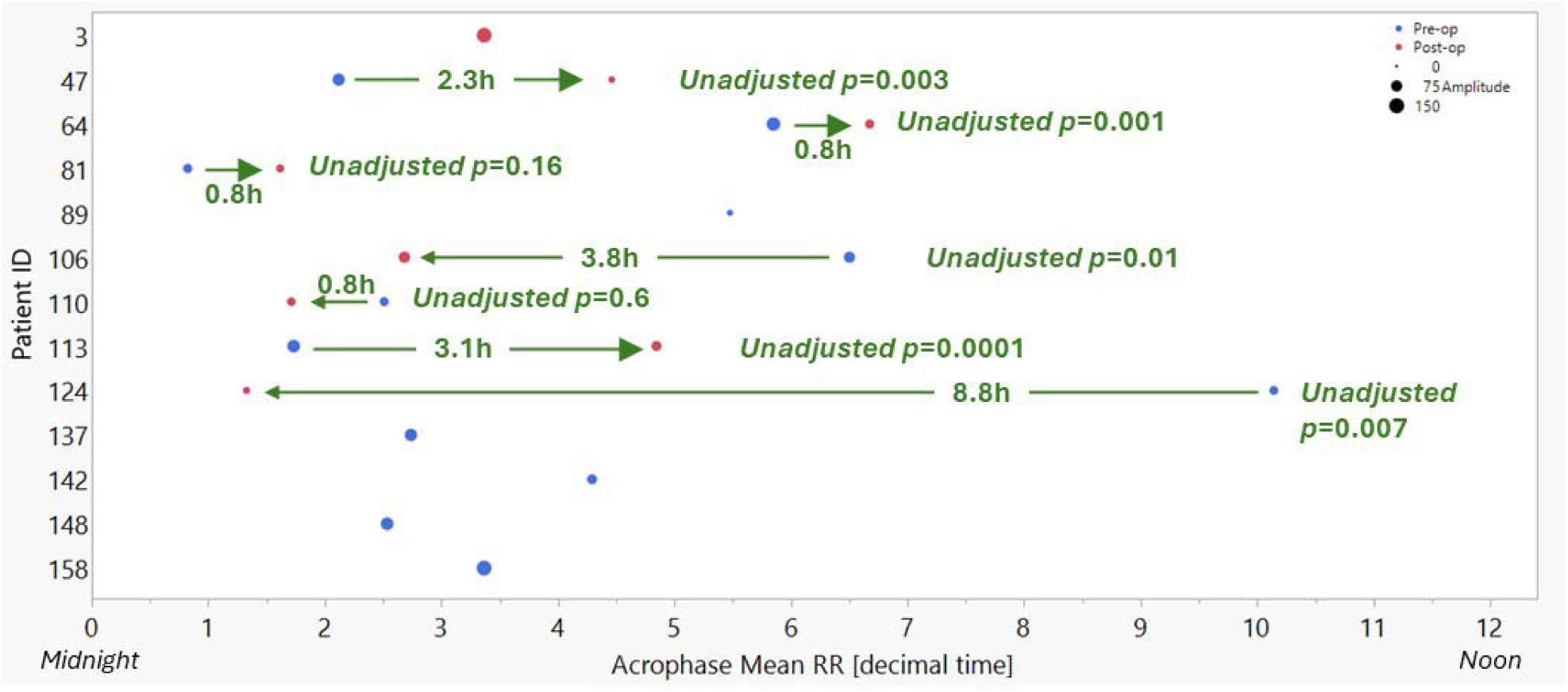
Acrophase-Shifts of RR Intervals in the SDU. Cosinor fits of ECG data collected with a chest-worn wearable device provided estimates for amplitude and acrophase of RR intervals as measure of heart rate variability. For seven patients the pre- to post-operative comparison of the acrophases suggested shifts of various lengths where significances observed in the unadjusted *p*-values can be conceptualized as experiencing acute jet lag in the intensive care environment. Note the directionality of the phase-shift indicating either phase advance, e.g. of 2.3h observed for #47, or phase delay, e.g. of 3.8h observed for #106. Size of dots indicates magnitude of RR interval amplitudes. Unadjusted *p*-values were derived from cosinor fits in the pre-op and post-op periods which were tested for differences in amplitude and phase for each individual by computing an F statistic using the linearHypothesis function from the car package in R (v3.1-3).

**Supplementary Figure 11.**
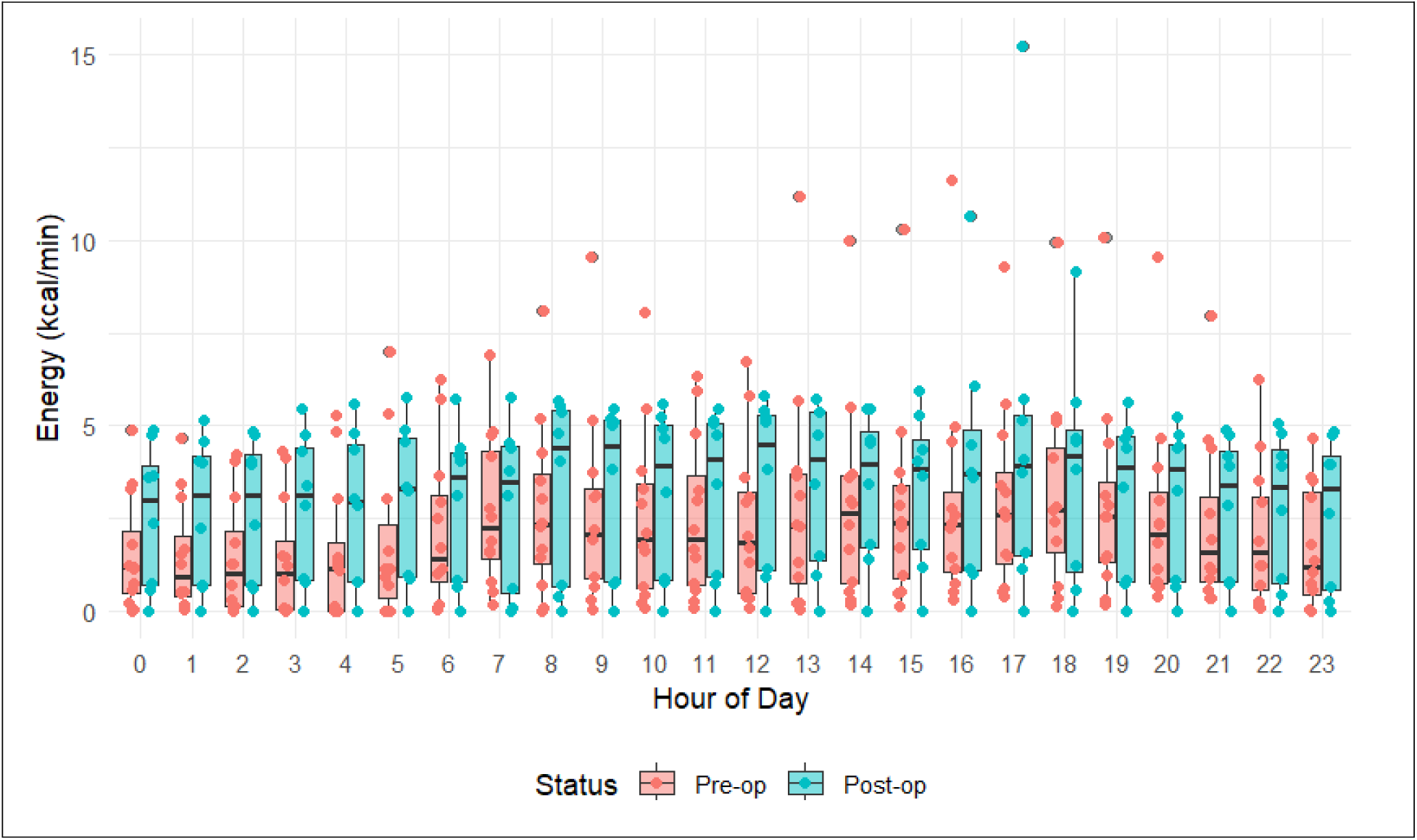
Metabolic Rate. The heart rate variability analysis of ECG wearable data produced estimates for metabolic rate plotted for the pre-operative and in-hospital conditions over 24 hours.

**Supplementary Figure 12.**
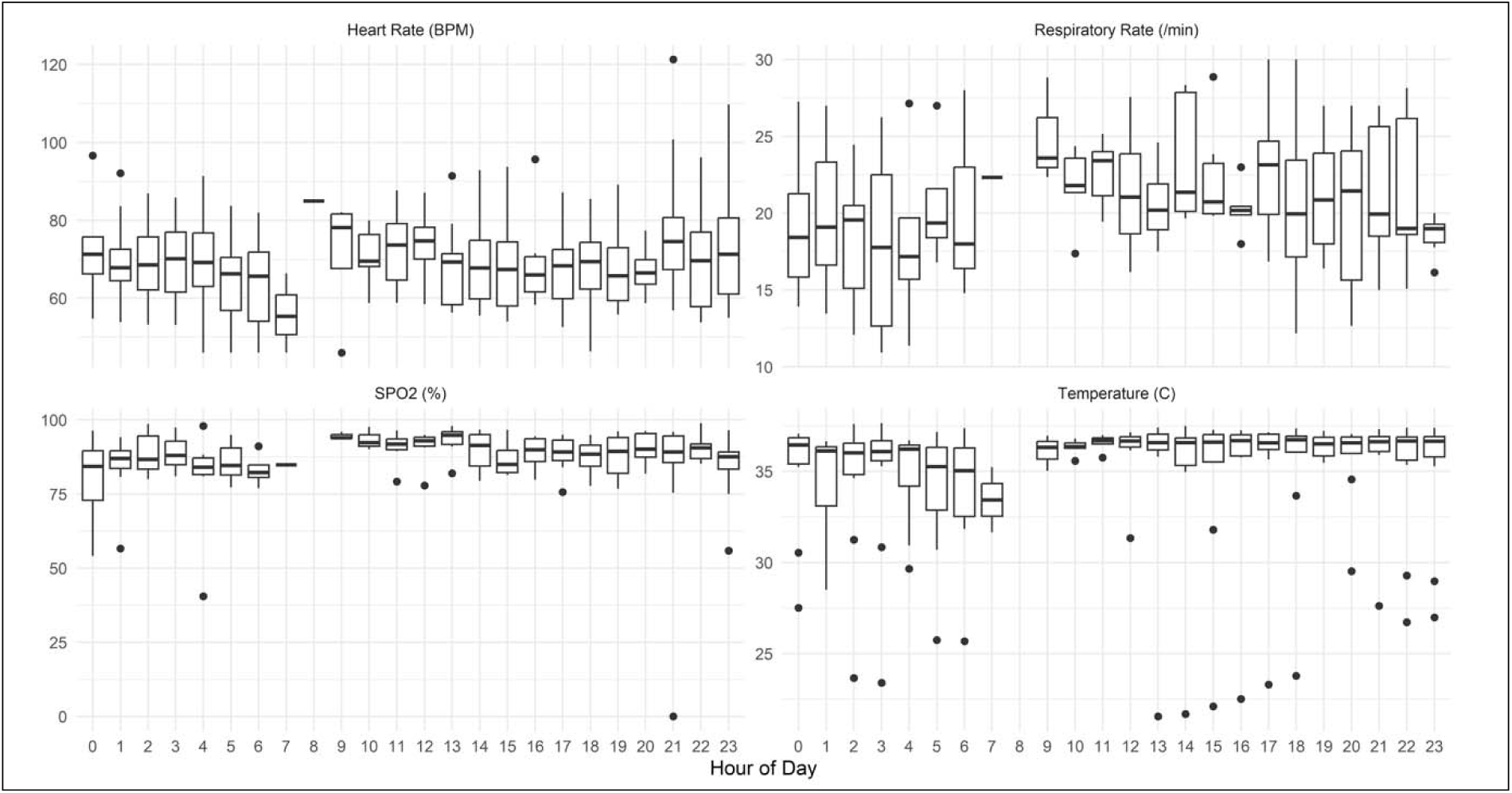
Vital Signs per In-Ear Wearable Device. A wearable device positioned into the external ear canal collected vital signs consisting of heart and respiratory rate, oxygen saturation and tympanic temperature.

**Supplementary Figure 13.**
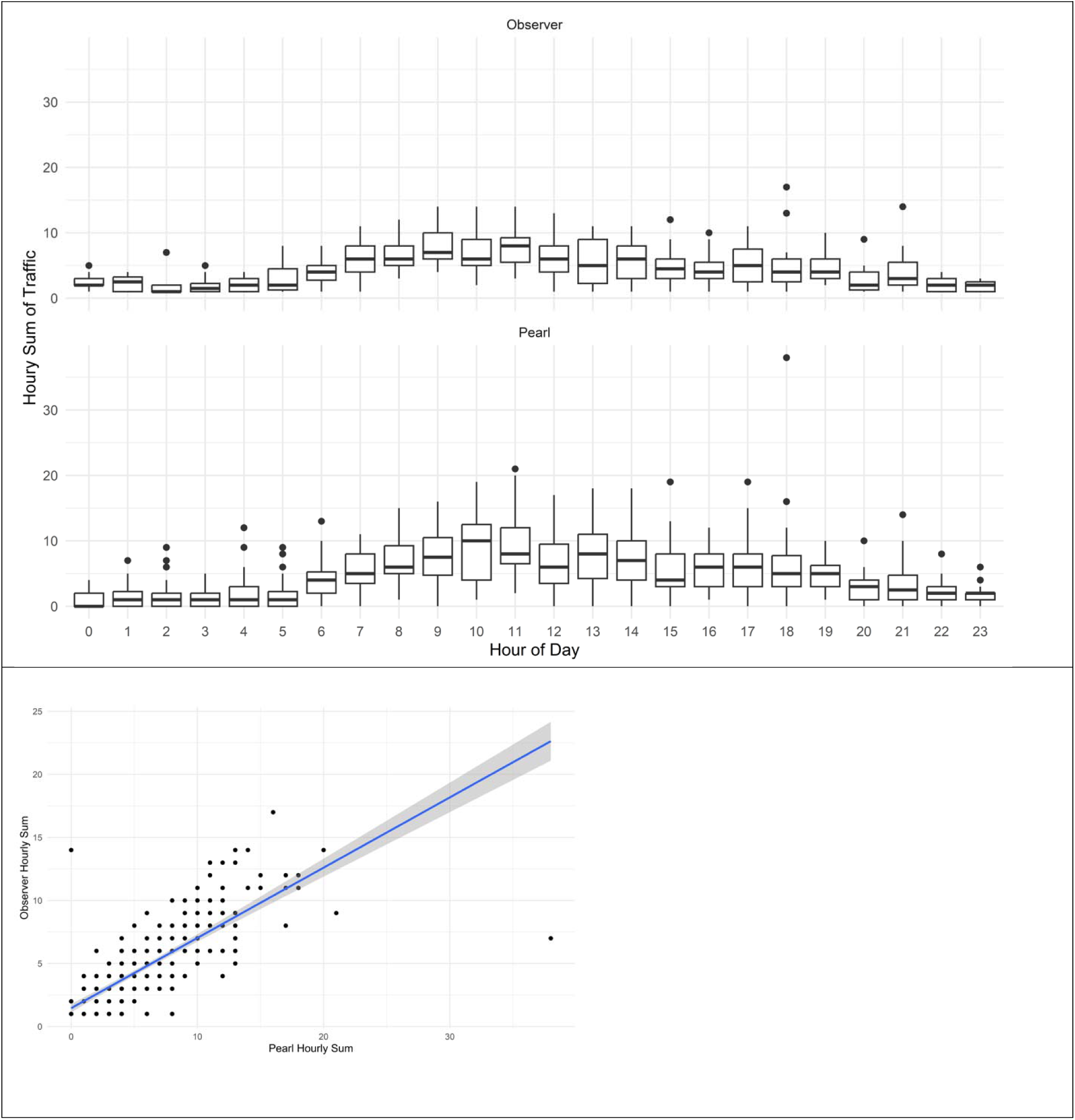
Observer & Sensor-Based Patient Room Traffic. Traffic into the patient rooms was logged with time stamps by student observers seated outside of the patient room (top) and sensors positioned at the door frame (center) which showed good agreement in the linear regression (bottom).

**Supplementary Figure 14.**
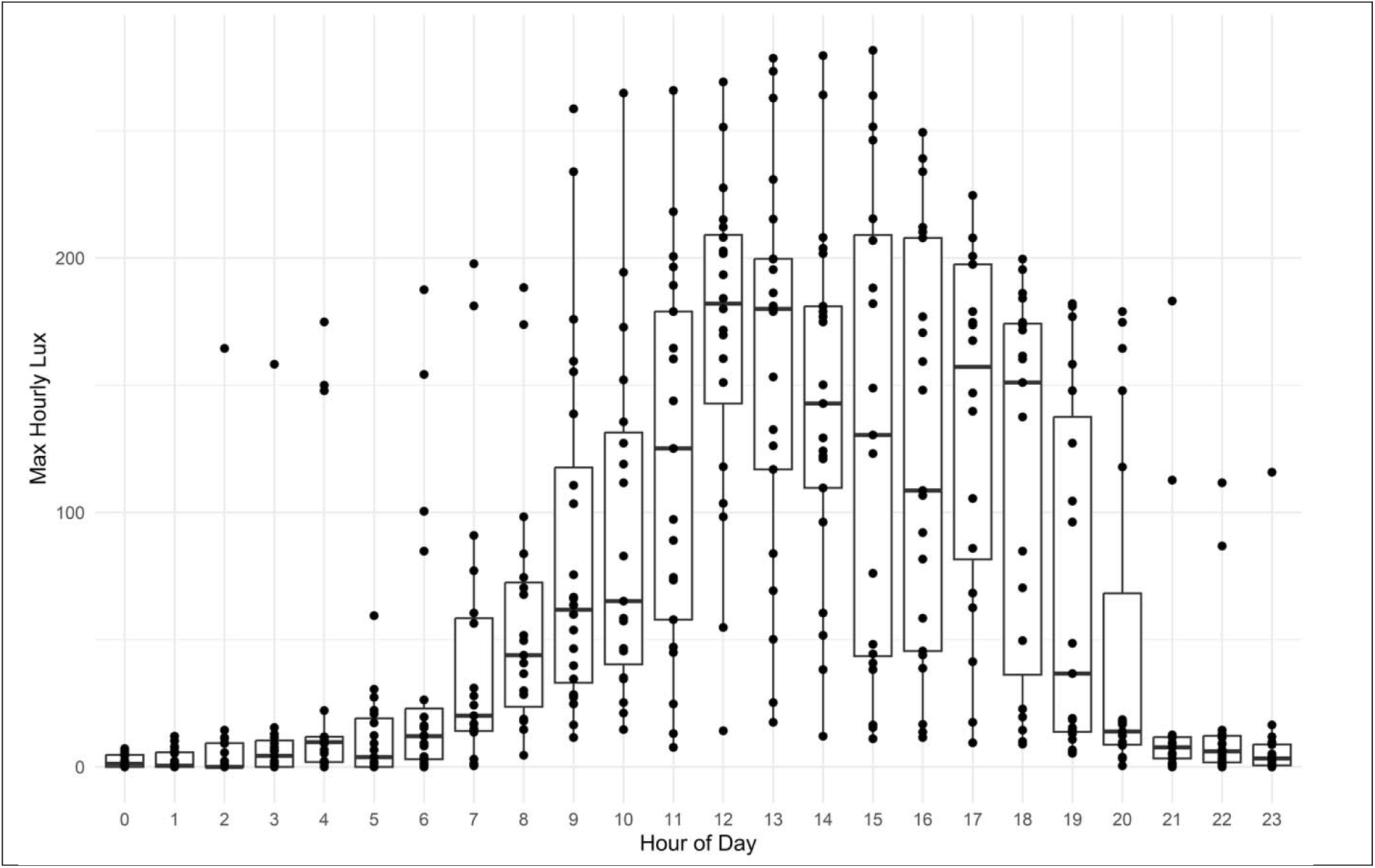
Luxmeter-Based Light Exposure. Diurnal traces of ambient light exposure in the patient room plotted for maximum hourly lux across the cohort.

**Supplementary Figure 15.**
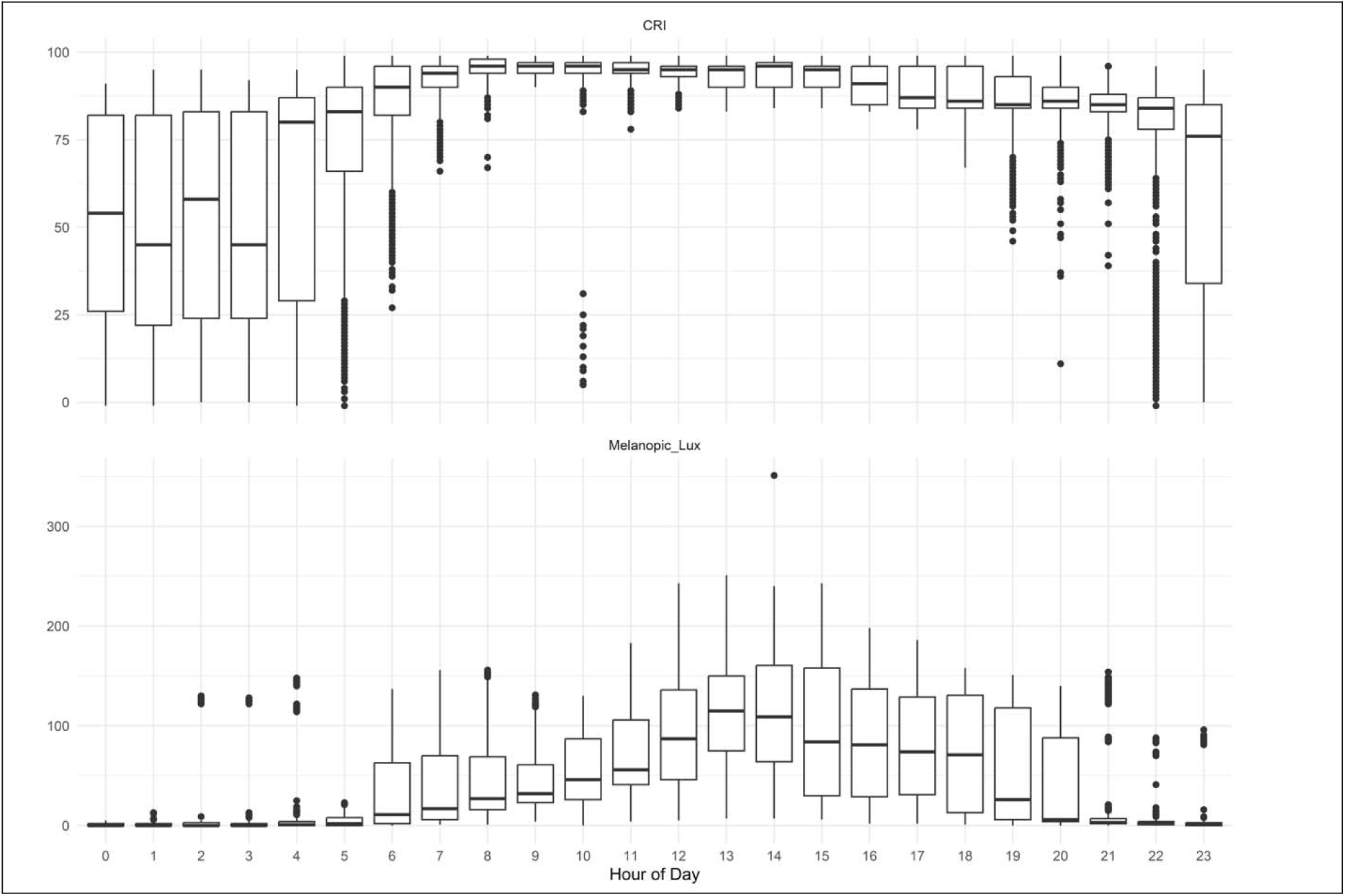
Color Rendering Index & Melanopsin Photoreception. A spectrometer device quantified the Color Rendering Index and melanopsin photoreception over 24 hours across the cohort.

**Supplementary Figure 16.**
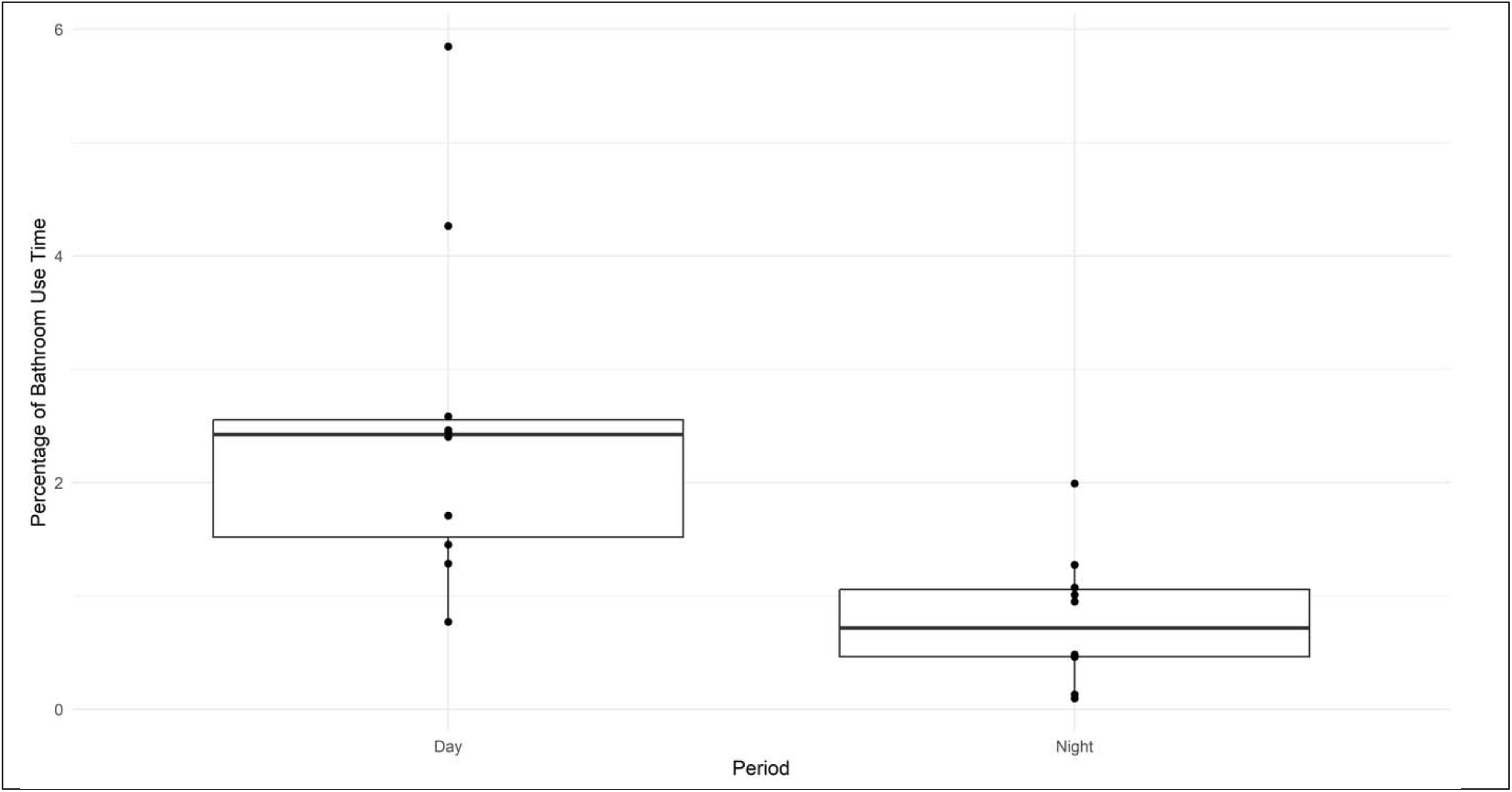
Bathroom Occupancy. A sensor mounted to the private bathroom ceiling collected occupancy data.

**Supplementary Figure 17.**
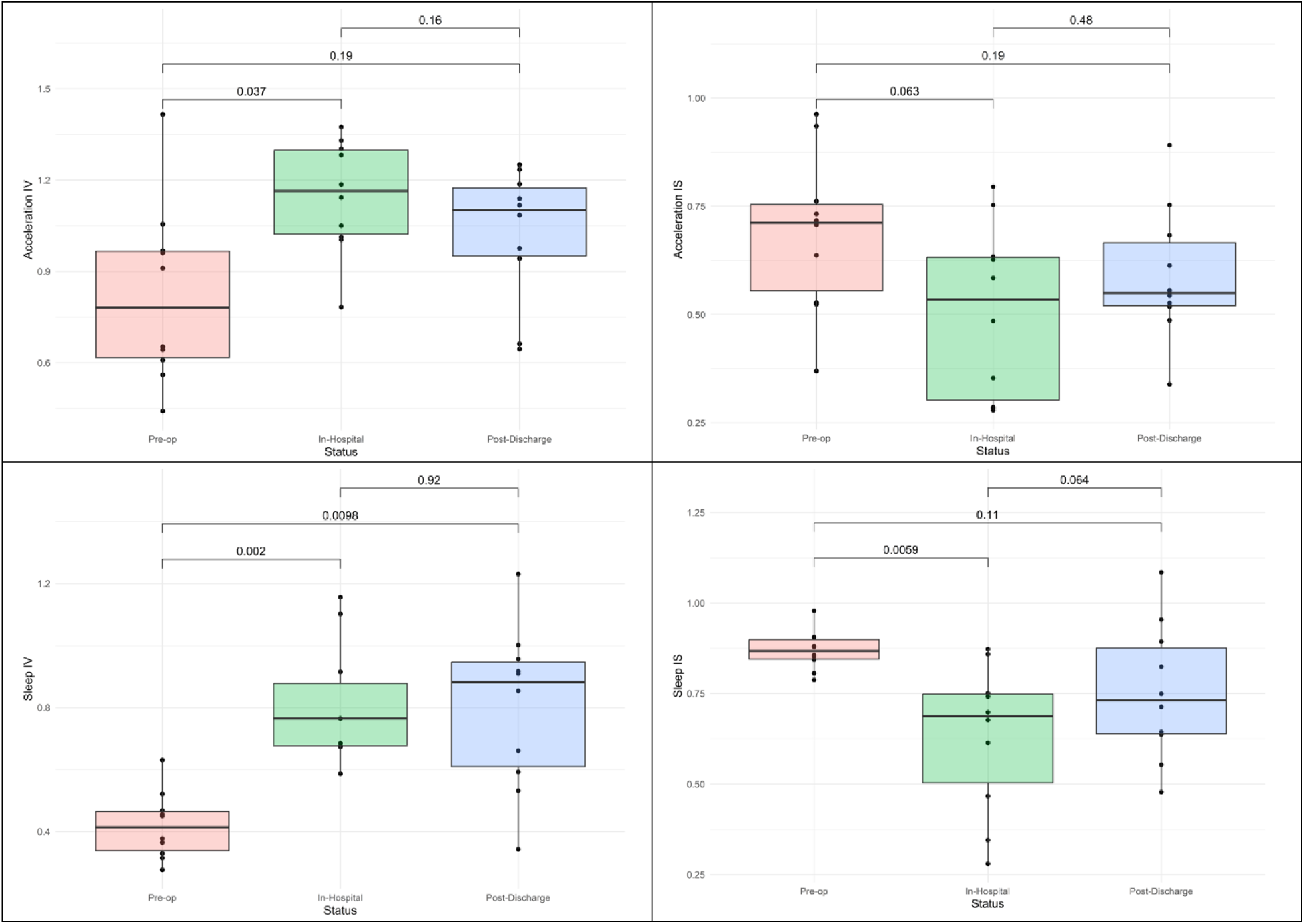
Robustness of Sleep-Wake Rhythms from Actigraphy. Intradaily variability and interdaily stability as measures of fragmented and robust daily rhythms, respectively, of physical activity and sleep derived from wrist accelerometry during the pre-operative, in-hospital and post-discharge conditions. P-values result from paired Wilcoxon tests.

**Supplementary Figure 18.**
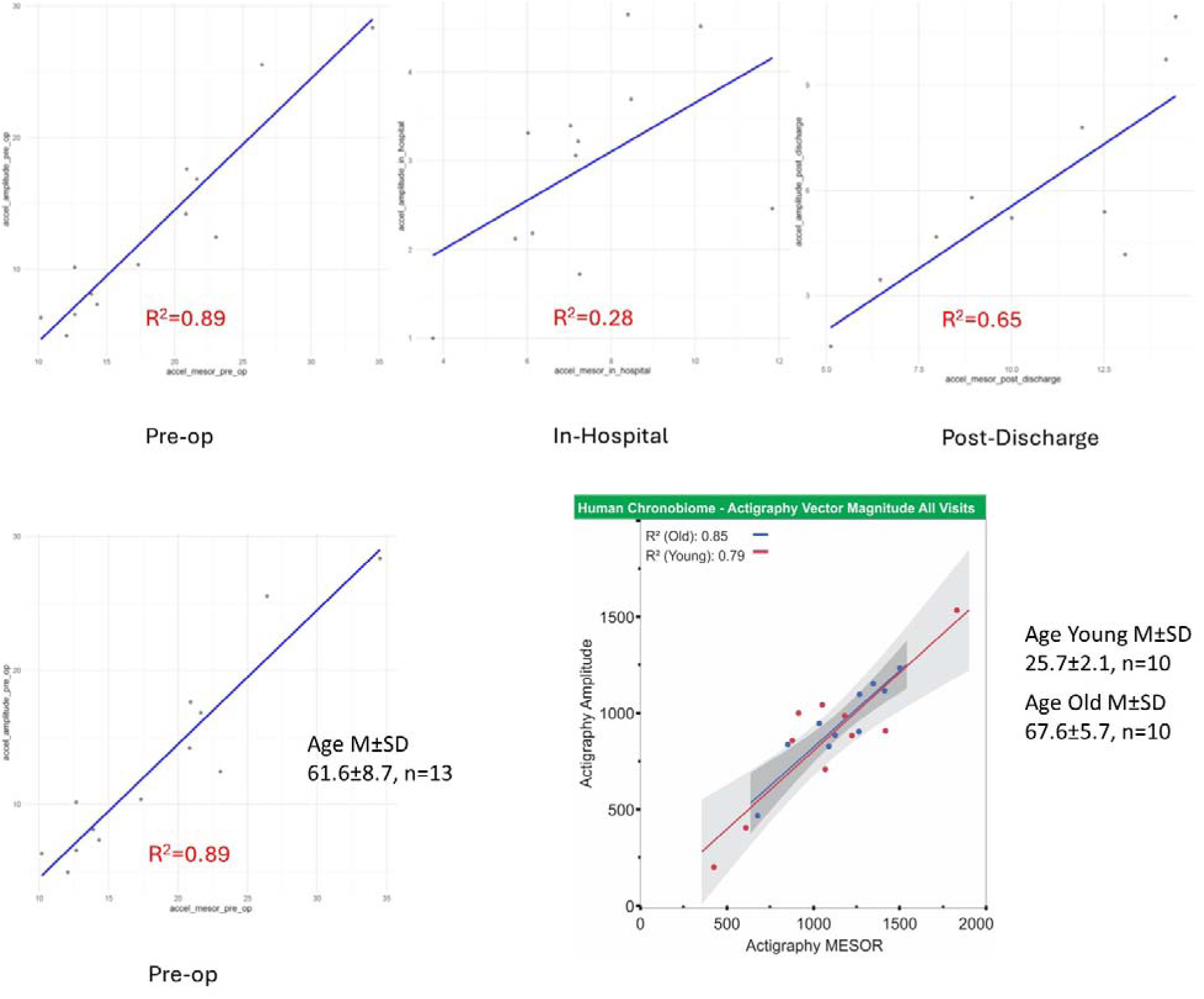
Correlation of MESOR & amplitude of physical activity. (Top) For physical activity, MESOR and amplitude estimates from the cosinor fits are correlated for the pre-operative, in-hospital and post-discharge conditions. R squared from linear regression. (Bottom) The difference in scale between the present study and the reference human chronobiome study is based on the difference in data outputs where the Axivity device reports g-force (present study) and the ActiGraph device in counts (Chronobiome study).

**Supplementary Figure 19.**
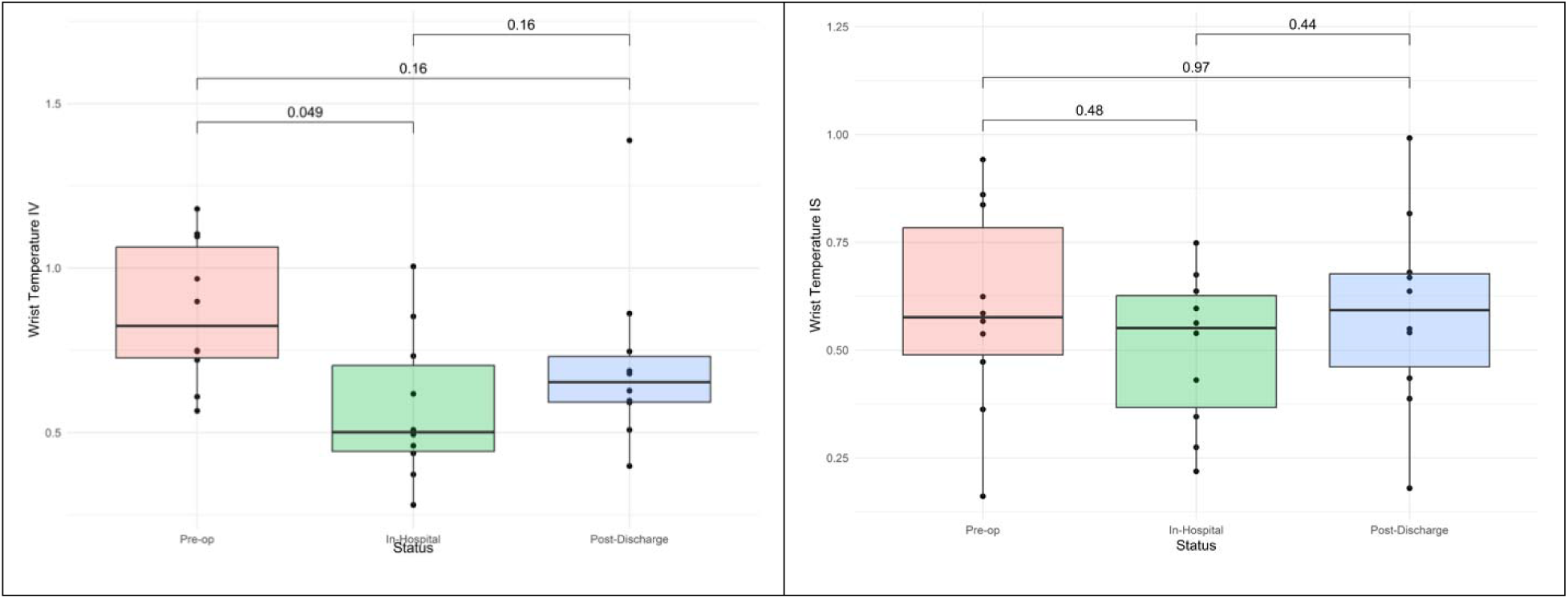
Robustness of Wrist Temperature Rhythms. Intradaily variability and interdaily stability as measures of fragmented and robust daily rhythms, respectively, of wrist temperature obtained from the wrist accelerometry device for the pre-operative, in-hospital and post-discharge conditions. P-values result from paired Wilcoxon tests.

**Supplementary Figure 20.**
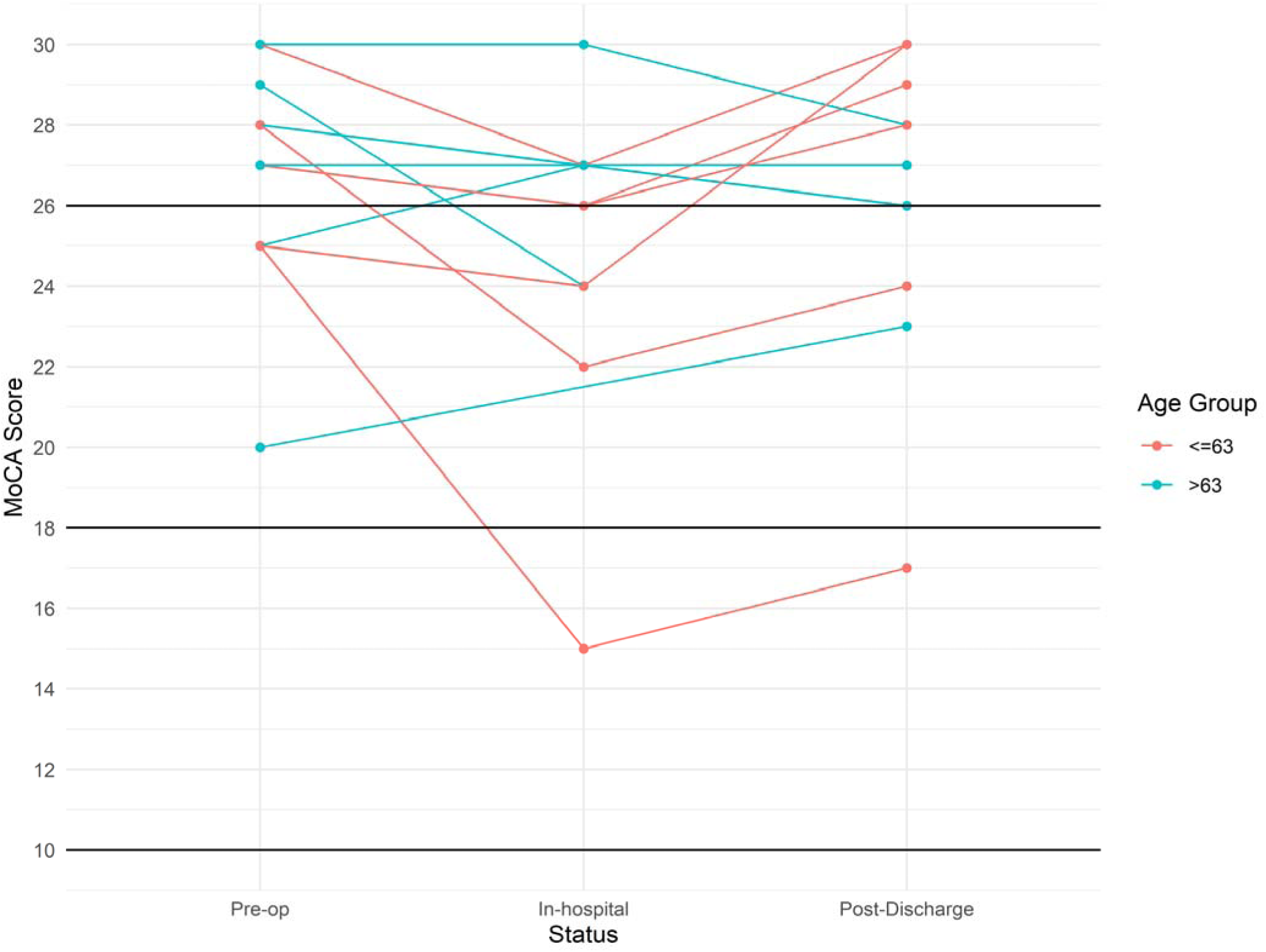
Patient-Level Trajectories of Cognitive Function. Cognitive function assessed by paper MoCA pre-operatively, in-hospital and post-discharge.

### Supplementary Tables

**Supplementary Table 1.** Medication Regimen in the SDU.

| ID | Anti-Arrhythmic & Betablocker | Antihypertensive & Diuretic | Sedative & Pain | Other Sedative & Sleep |
| --- | --- | --- | --- | --- |
| 3 | 1 | 1 | 0 | 0 |
| 47 | 1 | 1 | 1 | 0 |
| 64 | 0 | 1 | 1 | 0 |
| 81 | 0 | 1 | 1 | 0 |
| 89 | 1 | 1 | 1 | 1 |
| 106 | 1 | 0 | 1 | 0 |
| 110 | 1 | 1 | 1 | 0 |
| 113 | 0 | 1 | 1 | 1 |
| 124 | 1 | 0 | 1 | 0 |
| 137 | 1 | 1 | 1 | 0 |
| 142 | 1 | 1 | 1 | 0 |
| 148 | 1 | 1 | 1 | 0 |
| 158 | 1 | 1 | 1 | 0 |
| **Sum** | **10** | **11** | **12** | **2** |

Terms for “Anti-Arrhythmic & Betablocker” were amiodarone and metoprolol tartrate; for “Antihypertensive & Diuretic” amlodipine, furosemide, bumetanide, alfuzosin ER and hydrazaline; for “Sedative & Pain” tramadol, oxycodone and gabapentin; and for “Other Sedative & Sleep” melatonin and diphenhydramine.

**Supplementary Table 2.**
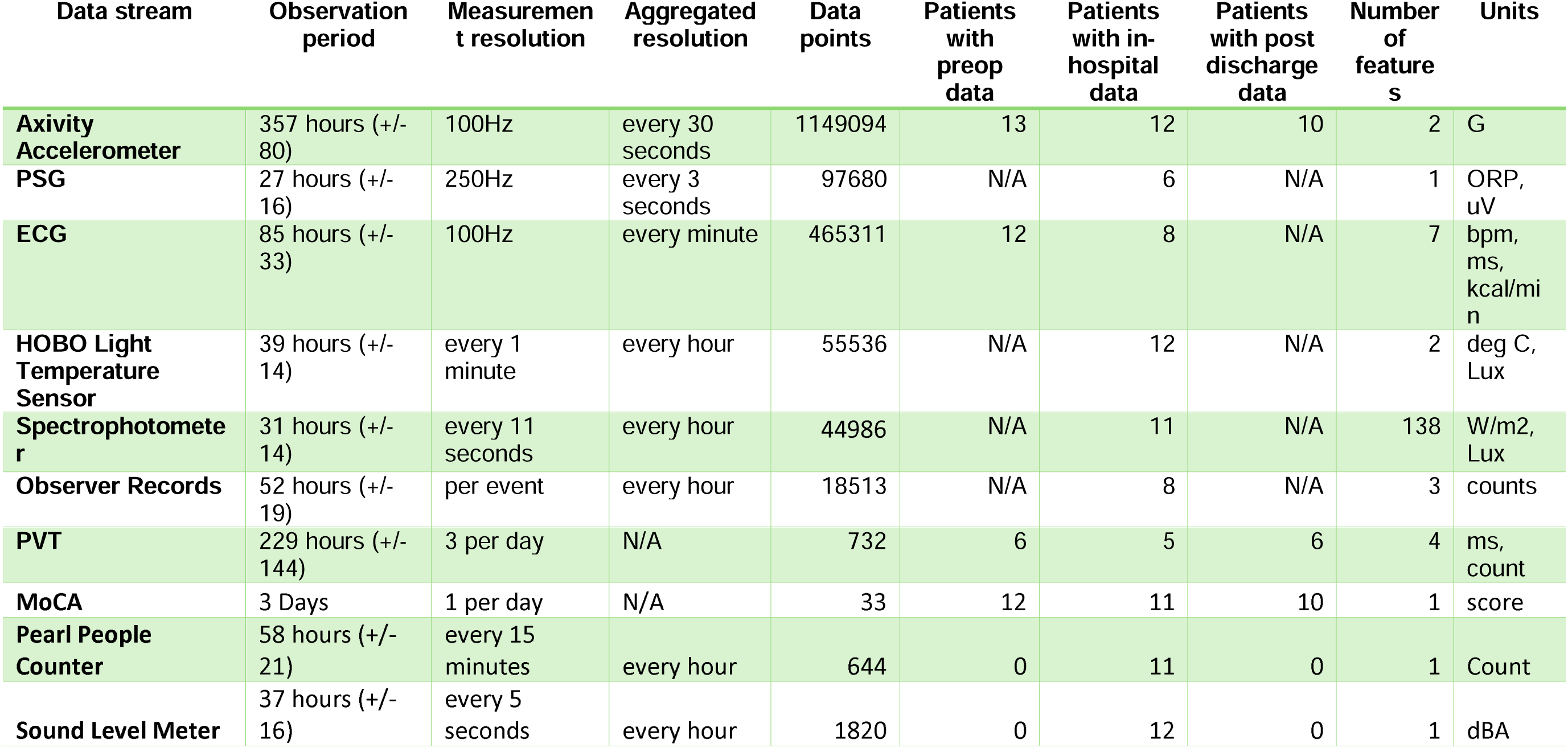
Number of Collected Data Points and Features, Sample Sizes and Units.

**Supplementary Table 3.** Emotional Valence.

| **Emotional**<br>**Valence** | **Pre-operative**<br>**Normalized**<br>**AUC** | **In-Hospital**<br>**Normalized**<br>**AUC** | **Post-Discharge**<br>**Normalized**<br>**AUC** | **Percent Change**<br>**Pre-op to In-**<br>**Hospital** | **Percent Change Pre-**<br>**op to Post Discharge** |
| --- | --- | --- | --- | --- | --- |
| *Angry* | 1.04 [0.10] | 1.37 [1.02] | 1.00 [0.21] | 31.8 | -3.7 |
| *Anxious* | 1.75 [2.11] | 3.99 [2.46] | 1.93 [0.69] | 127.5 | 10.0 |
| *Blue* | 1.09 [0.62] | 1.68 [2.63] | 1.42 [1.31] | 53.5 | 29.7 |
| *Energetic* | 4.56 [2.05] | 3.28 [1.38] | 4.02 [0.44] | -28.1 | -11.9 |
| *Frightened* | 1.45 [1.59] | 2.37 [2.57] | 1.40 [0.64] | 64.0 | -3.1 |
| *Happy* | 7.94 [0.54] | 5.29 [1.08] | 5.88 [2.32] | -33.4 | -26.0 |
| *Hopeful* | 7.79 [1.37] | 5.62 [1.85] | 6.89 [2.22] | -27.9 | -11.6 |
| *Irritable* | 1.55 [0.73] | 1.73 [2.12] | 1.54 [0.49] | 11.4 | -0.4 |
| *Lonely* | 1.04 [1.57] | 1.14 [1.99] | 1.00 [1.01] | 8.8 | -4.2 |
| *Sleep Quality* | 1.92 [0.72] | 1.65 [0.35] | 1.92 [0.56] | -14.1 | 0.0 |

**Supplementary Table 4.** Wrist Temperature.

| Measurement | Median<br>Pre-op | Median In-Hospital | Median<br>Post-Discharge | p-value<br>(Pre-op - In-Hospital) | p-value<br>(Pre-op<br>Post-Discharge) | p-value<br>(In-Hospital<br>Post-Discharge) |
| --- | --- | --- | --- | --- | --- | --- |
| MESOR | 20.6 [0.7] | 20.2 [0.9] | 20.4 [1.3] | 0.2 | 0.7 | 0.0 |
| Amplitude | 0.7 [0.5] | 0.6 [0.3] | 0.7 [0.4] | 0.3 | 0.4 | 0.6 |
| Phase | 2.2 [13.3] | 1.9 [2.0] | 1.6 [1.3] | 0.7 | 0.6 | 0.4 |
| Interdaily<br>Stability | 0.6 [0.3] | 0.6 [0.3] | 0.6 [0.2] | 0.3 | 0.6 | 0.5 |
| Intradaily<br>Variability | 0.8 [0.3] | 0.5 [0.3] | 0.6 [0.1] | 0.0 | 0.2 | 0.2 |

**Supplementary Table 5.** ECG and Heart Rate Variability.

| Index | Condition | Median [IQR]<br>Amplitude | Amplitude p-value | Median [IQR]<br>MESOR | MESOR p-value |
| --- | --- | --- | --- | --- | --- |
| Heart Rate | Pre-op | 4.64 [3.19] | Ref | 67.8 [6.4] | Ref |
| Heart Rate | Post-op | 4.05 [2.59] | 0.937 | 78.7 [10.5] | 0.078 |
| PNS Index | Pre-op | 0.46 [0.28] | Ref | -0.7 [0.5] | Ref |
| PNS Index | Post-op | 0.33 [0.40] | 0.938 | -1.3 [1.0] | 0.297 |
| RMSSD | Pre-op | 3.22 [1.67] | Ref | 20.7 [4.5] | Ref |
| RMSSD | Post-op | 1.72 [13.85] | 0.937 | 13.7 [18.6] | 0.937 |
| RR Interval | Pre-op | 74.7 [52.6] | Ref | 897.4 [74.6] | Ref |
| RR Interval | Post-op | 39.1 [23.9] | 0.031 | 765.2 [100.7] | 0.078 |
| SDNN | Pre-op | 2.6 [2.34] | Ref | 22.3 [6.5] | Ref |
| SDNN | Post-op | 2.24 [7.52] | 0.938 | 10.8 [13.0] | 0.219 |
| SNS Index | Pre-op | 0.67 [0.40] | Ref | 1.3 [1.1] | Ref |
| SNS Index | Post-op | 0.79 [0.34] | 0.156 | 4.1 [3.1] | 0.031 |

**Supplementary Table 6.** In-Ear Vital Signs.

| Measurement (Cosinuss device) | Rhythm Metric | Median | IQR |
| --- | --- | --- | --- |
| Heart Rate | Amplitude | 4.9 | 5.7 |
| Heart Rate | MESOR | 73.2 | 8.0 |
| Heart Rate | Phase | 3.9 | 15.0 |
| Respiratory Rate | Amplitude | 5.0 | 4.2 |
| Respiratory Rate | MESOR | 19.5 | 4.7 |
| Respiratory Rate | Phase | 14.1 | 3.3 |
| SpO2 | Amplitude | 5.0 | 14.6 |
| SpO2 | MESOR | 84.1 | 4.6 |
| SpO2 | Phase | 11.4 | 13.9 |
| Temperature | Amplitude | 0.9 | 2.2 |
| Temperature | MESOR | 36.0 | 1.2 |
| Temperature | Phase | 17.2 | 3.6 |

**Supplementary Table 7.** Deployment and Compliance of PSG and In-Ear Vital Signs Device.

| Patient ID# | Cerebra Device Placed | Cerebra Device Sliding | Cerebra Device Declined due to other reason | Cerebra Device Not Placed due to logistical reason | Cosinuss Device Placed | Cosinuss Device Dislodged | Cosinuss Device Declined due to other reason | Cosinuss Device Not Placed due to logistical reason |
| --- | --- | --- | --- | --- | --- | --- | --- | --- |
| 3 | 1 | 1 | 0 | 0 | 1 | 0 | 0 | 0 |
| 47 | 1 | 1 | 0 | 0 | 1 | 1 | 0 | 0 |
| 64 | 1 | 0 | 1 | 0 | 0 | 0 | 1 | 0 |
| 81 | 1 | 0 | 0 | 0 | 1 | 0 | 0 | 0 |
| 89 | 0 | 0 | 0 | 1 | 0 | 0 | 0 | 1 |
| 106 | 0 | 0 | 1 | 0 | 0 | 0 | 1 | 0 |
| 110 | 0 | 0 | 1 | 0 | 1 | 0 | 0 | 0 |
| 113 | 1 | 1 | 0 | 0 | 1 | 0 | 0 | 0 |
| 124 | 0 | 0 | 1 | 0 | 1 | 0 | 1 | 0 |
| 137 | 0 | 0 | 1 | 0 | 0 | 0 | 1 | 0 |
| 142 | 0 | 0 | 1 | 0 | 1 | 0 | 0 | 0 |
| 148 | 0 | 0 | 0 | 1 | 1 | 0 | 0 | 0 |
| 158 | 1 | 0 | 1 | 0 | 1 | 0 | 0 | 0 |
| **Sum** | **6** | **3** | **7** | **2** | **9** | **1** | **4** | **1** |

## Notes

### Competing Interest Statement

The authors have declared no competing interest.

### Clinical Trial

NCT05828680

### Author Declarations

The Institutional Review Board (IRB) of the University of Pennsylvania gave ethical approval for this work

